# Estimating the causal effect of body mass index on gut microbiota variation

**DOI:** 10.1101/2025.08.18.25333926

**Authors:** David A. Hughes, Laura J. Corbin, Sebastian Proost, Kaitlin H. Wade, Jeroen Raes, Nicholas J. Timpson

## Abstract

The gut microbiome is a complex ecosystem which has population-level variation in composition and abundance known to be associated with body mass index (BMI). The nature of these relationships is largely unknown and potentially confounded by environment, diet and other behavioral traits. We used fecal 16S rRNA data from 2,225 individuals from the Flemish Gut Flora project to derive 368 microbiota traits (MTs) including diversity, abundance, presence, or absence, enterotype class and abundance ratio. We assessed the relationship between BMI and those traits in observational and Mendelian randomization (MR) frameworks, the latter being used to estimate a causal effect of BMI on MT variation. We found that 141 (38%) of MTs associated with BMI in observational analyses. These estimates showed concordance with those from MR, suggesting that BMI has a broad influence on gut MTs. Evidence here was strong enough to suggest causal BMI effects for 41 MTs (11% of all those tested). This included a reduction in overall diversity, a decrease in the abundance of genera *Barnesiella* of phylum Bacteroidetes (B) and a decrease in the abundance of genera *Sporobacter* of phylum Firmicutes (F) with higher BMI. There was also an increase in the ratio of genera *Roseburia* to genera *Sporobacter* (F/F), genera *Oscillibacter* to *Sporobacter* (F/F), genera *Bacteroides* to *Sporobacter* (B/F), and *Bacteroides* to *Barnesiella* (B/B) with increases in BMI. The log-odds of assignment to enterotype Bacteroides2 increased and – in females – the presence of genera Adlercreutzia decreased with higher BMI. Previous studies have implied that variation in the gut microbiome influences BMI, but the present results support the conclusion that relationships between BMI and microbiome variation are, at the very least, bi-directional.

## Introduction

Rates of overweight and obesity are increasing globally, and this is clearly escalating the overall burden of ill-health [1]. Body mass index (BMI), or the ratio of an individual’s weight and height squared (kg/m^2^), is an imperfect but common and useful measure of excess weight or fat and a metric used to assign individuals to discrete adiposity-related health and disease categories, namely underweight, healthy, overweight and obesity [2–5]. Cross-sectional increases in BMI have been associated with a variety of diseases including cardiometabolic disease, cardiovascular disease and type 2 diabetes [6], as well as with some cancers [7–9] and decreased life expectancy [10–13].

Microorganisms are a potentially important part of symbiotic life that could influence host phenotype [14–16]. Numerous studies have illustrated that gut microbiota are associated with environmental and host physiology variables [17–23]. Notably, these associations extend to variation in BMI and a wide range of diseases, including metabolic disorders such as type 2 diabetes, inflammatory bowel syndrome, Crohn’s disease, neurological traits such as Parkinson’s disease and stress, as well as cancers and other complex traits [24–37]. Previous work in mice and humans has illustrated that the abundance of microbiota taxa varies between lean individuals and those with obesity [38]. Further, changes in weight or BMI through diet modulates microbiota taxa abundances, suggesting that BMI can alter variation in microbiota abundances [39]. In a potentially reciprocal manner, the introduction of gut microbiota derived from lean mice into germ free mice increases body fat and the introduction of gut microbiota from leptin-deficient obese mice is associated with further increases in body fat [40], suggesting that altering the gut microbiota could modulate variation in BMI. Whilst similar efforts to change the gut microbiota in humans have not resulted in evidence for causal changes in BMI [41], there is evidence suggesting that therapeutic and dietary interventions – which alter gut microbiota – are in turn associated with improvement in metabolic traits and weight [23,42,43]. Additionally, early life exposure to antibiotics, which is thought to alter gut microbiome composition, increases BMI within the first 7 years of life [44]. With this, there have been suggestions that the microbiome composition and function may influence energy harvesting from the diet and energy storage in the host [45], acting through a variety of mechanisms [46].

Common difficulties with observational analysis of variation in gut microbiota and disease traits are confounding and reverse causality [47]. Specifically, distinguishing whether variations in gut microbiota precede and influence disease risk, are caused by the disease itself, or simply coincide with disease due to shared confounders (such as diet, lifestyle, or genotype) is difficult to ascertain without well-executed prospective studies and randomized controlled trials. In lieu of these studies, Mendelian randomization (MR) may aid assertions of causality in studies of microbiome and BMI [48,49]. MR is a genetic epidemiology approach that promotes the use of genetic variation associated with modifiable exposures to aid the assertion of causality. In the context of this work, gut microbiota are modelled as a response variable within a MR framework to assess the evidence for causal effects of BMI on microbial variation (**Figure 1**). The advantage to this approach is that genotypes used as instruments for our exposure trait (BMI) are robustly associated with BMI but only reflect variation which is theoretically free from more conventional confounding, and which occurs upstream of observed outcomes (i.e., avoiding reverse causation). Assessing this direction of effect is of particular interest considering the substantial observational literature that is unable to make inferences of directionality and given the lack of microbiome effect estimates from studies of change or variation in BMI [50–59].

**Figure 1.**
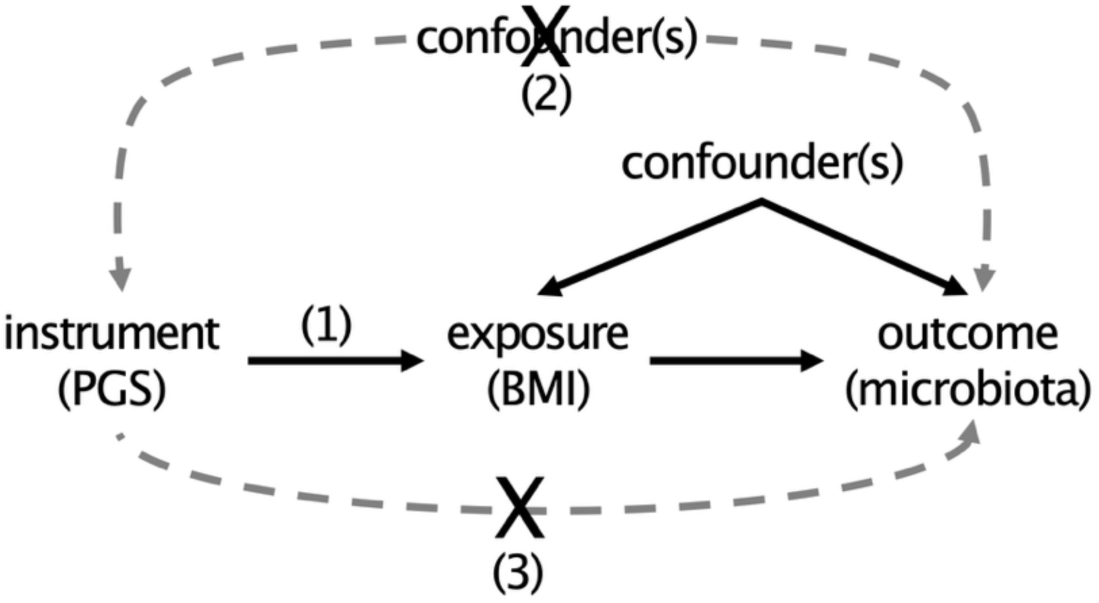
Mendelian Randomization DAG. A schematic representation of a directed acyclic graph (DAG) of the Mendelian randomization framework. The MR framework in this study assumes (1: relevance) that the instrument (PGS) is robustly associated with the exposure (BMI), (2: independence) that there are no confounders of the instrument outcome (microbiota) association and (3: exclusion restriction) that the instrument acts on the outcome only through the exposure and not through any pleiotropic pathway.

Whilst it would be ideal to also test the reverse (i.e., consider the gut microbiota as the exposure), these analyses are fraught with difficulties [60,61]. First and foremost, only a single robust association between host genotype and gut microbial variation has been identified to date [60,62–66]. Namely, the association between *Bifidobacterium* and the lactose tolerance allele (rs4988235) at the MCM6/LCT locus. However, this variant influences multiple factors, including host behavior and environment, and has numerous pleiotropic associations, making it difficult to infer a direct causal path from exposure (*Bifidobacterium*) to any outcome of interest [60,61].

We aimed to provide two estimates of the association between BMI and gut microbiota variation. First, human genetic and 16S rRNA gut microbiota data from the Flemish Gut Flora Project (FGFP) were used to estimate a conventional cross-sectional estimate of the effect of BMI on variation in fecal microbiota. Second, a within-sample or one-sample MR framework [67] was used to obtain a causal effect estimate of BMI on variation in gut microbiota. Analyses were conducted in a sex-combined and sex-stratified analyses – motivated by the dimorphism in adiposity distributions among the sexes [68], the large associated contribution of sex to variation in the gut microbiota [18], and variation in disease risk among the sexes [69,70]. In the MR analysis, we used an instrument that is robustly associated with BMI to provide an estimate of the direct causal effect of BMI on gut microbiota variation.

## Materials and methods

### Study sample

The study sample population was from the Flemish Gut Flora Project (FGFP), a population-based study in the Flanders region of Belgium focused on gut flora and health [18]. Sampling procedures are described in full elsewhere [18,60]. Volunteers were invited to participate through local media and provided informed consent by mail. FGFP procedures were approved by the Medical Ethics Committee of the University of Brussels/Brussels University Hospital (approval no. 143201215505, 5 December 2012) and a declaration concerning the FGFP privacy policy was submitted to the Belgian Commission for the Protection of Privacy. Between June 2013 and April 2016, medical questionnaires were completed by each volunteer’s general practitioner who also recorded anthropometric and basic clinical measurements. Sex was self-reported. Stool samples were collected remotely with sampling kits sent to volunteer homes. Samples were collected and stored by volunteers at -18°C. Upon arrival at the research facility, they were stored again at -18°C until collection and long-term storage at -80°C.

Data from general practitioners included drug use in the form of Anatomical Therapeutic Chemical or ATC codes. To account for antibiotic drug use in our analyses, we grouped drugs into their higher order categories [71,72]. The three categories used here were J01 antibacterials (n=617), A07A intestinal anti-infective (n=2), and S01A or ophthalmological anti-infective (n=24). In addition, we identified those individuals on A07 anti-diarrheal (n=94), and A07 anti-constipation medicines (n=31).

### Microbiome data

Hypervariable region V4 of the 16S rRNA gene was amplified from DNA derived from frozen fecal samples collected from 2,482 FGFP volunteers using the 515F/806R primer pair (GTGYCAGCMGCCGCGGTAA and GGACTACNVGGGTWTCTAAT) modified to include barcodes to identify each sample [73]. Libraries were sequenced on the Illumina HiSeq platform at the VIB Nucleomics core laboratory (Leuven, Belgium) and fastq sequences were phylogenetically assigned using the DADA2 pipeline (v.1.6) with rarefied data at 10,000 reads per sample [74]. Complete details of DNA preparation, library preparation, sequencing and data processing can be found in Hughes *et al*. [60].

### Defining microbial traits

Derivation of MTs follows procedures outlined previously [60]. In brief, rarefied (i.e., sub-sampled to a uniform read depth) DADA2 count data was used to define five types of MT: diversity (1. Div) including both α-diversity (i.e., within-person) and β-diversity (i.e., between-person) metrics; Enterotype (2. Entero); abundance (3. AB) traits including zero-truncated abundance (trunc_AB; where all zero count values were removed); binary presence/absence (4. PA) traits; and ratios (5. RA) derived from the ratio of correlated AB traits (**Supplementary Figure 1**). Taxa labeled as ‘unclassified’ represent 16S rRNA sequence variants that could be assigned to a higher-level taxonomic rank (e.g., phylum or family) but not to a more specific lower-level rank (e.g., genus), as determined by the DADA2 taxonomic assignment pipeline. α- and β-diversity metrics were estimated using the DADA2 rarefied data and 288 genus-level counts, using the ‘vegan’ R package [75]. α-diversity metrics (n = 3) include the number of genera, Shannon diversity, and Chao diversity. β-diversity (n = 2) was derived from a two-axis non-metric multi-dimensional scaling analysis. Four enterotypes were defined using Dirichlet multinomial mixtures [76] using the ‘DirichletMultinomial’ R package [77]. Enterotypes were then transformed into binary Entero (n = 4) traits defined as being (1) or not-being (0) assigned to each of the four enterotypes.

PA traits were generated for all phylogenetic units (taxa) that had a minimum of 250 present (non-zero) and 250 absent (zero) observations (n = 156 PA MTs). A trunc_AB trait was generated for all taxa that had a minimum of 1000 present observations, a minimum of 250 absent observations, and a zero-truncated mean abundance of 20 or greater (n = 55 trunc_AB MTs). All zero values in trunc_AB traits were turned into NAs and, as a result, trunc_AB varied in sample size across MTs.

AB traits, in contrast, retain all zero observations and were generated for all taxa that have non-zero observations for at least 75% of samples, and a zero-truncated mean abundance of 20 or greater (n = 73 AB MTs). Thresholds used to define PA and AB traits (e.g., minimum numbers of present and absent observations or non-zero observations) were selected to balance statistical power and trait count burden. These PA, trunc_AB, and AB MTs (n = 284 MTs) were built using the bespoke R function build_mts(), which can be found in the study GitHub repository [78]. MT names include in the suffix a numerical value, such as 0.433, as seen in G_Methanobrevibacter_0.443_trunc_AB. This value represents the proportion of the sample population that had non-zero values for this microbial taxon and provides the reader with a quick and easy evaluation of how prevalent the trait is in the sample population. Zero truncated traits retain a trunc_AB suffix in their names but, in tables and figures, they will be included in the AB grouping.

RA traits were built using AB traits, no trunc_AB traits were included. A Spearman’s correlation coefficient was estimated among all AB traits with a median count of at least 25, and all AB trait pairs with an absolute rho greater than or equal to 0.2 and smaller than or equal to 0.9 were used to build RA traits. The defined correlation thresholds were used to ensure that ratio traits were constructed from taxa pairs with sufficient co-variation to plausibly reflect coordinated ecological adaptation, while also limiting the overall number of traits derived.

The defined MTs (n=815; Div = 5 (α-diversity = 3), Entero = 4, AB = 128 (trunc_AB = 55), PA = 156, RA = 522; **Supplementary Table 1**) contained correlated and redundant (identical values) MTs (**Supplementary Figure 2**). To define a primary analysis set of traits that excludes highly correlated or redundant traits, a Spearman’s rho (using pair wise complete observations) was estimated between all pairs, continuous and binary, of AB, RA, and PA MTs (**Supplementary Table 2**). Note that we chose to retain all Div and Entero MTs as primary traits and excluded them from this analysis. Then, a distance matrix was constructed where the distance was defined as one minus the absolute value of Spearman’s rho, a hierarchical clustering dendrogram was constructed using the hclust() function and the method “complete” from the R stats package and finally a tree cut was performed using the cutree() function from the stats package using a tree cut height (parameter “h”) of 0.05. This cut height threshold (h) identifies clusters of traits with a Spearman’s rho greater that 0.95 allowing us to remove, what we deem to be, statistically redundant traits. In addition, we favored lower order taxonomic MTs of the same trait type (AB, trunc_AB, PA, RA) over higher order taxonomic MTs in the list. In total, 359 MTs (AB = 67, PA = 104, RA = 188) remained in our primary analysis MT set and 447 MTs were removed. When combined with the three α-diversity metrics, two β-diversity and four binary enterotype traits, this totaled 368 MTs in the primary analysis set (**Supplementary Figure 3**). All analytical results for all 815 MTs are available in **Supplementary Materials** but only results from the (non-redundant) primary analysis set (n = 368) will be presented and discussed.

### Effective number of microbial traits

To determine the effective or independent number of MTs that remain in this inter-correlated data set (n = 368, **Supplementary Figure 3**), we followed the procedure of Gao *et al*. [79]. First a Spearman’s correlation matrix was generated among all MTs in the primary data set. Second, a principal component analysis was performed using this correlation matrix, which allows for data missingness and has the advantage of not requiring a data transformation given the use of a ranked correlation (**Supplementary Table 2, Supplementary Figure 4**). Third, we identified the number of components needed to explain at least 95% of the total variance in the data set. From this, we estimated that there are 33 effectively independent MTs in the data set. This estimate of the effective number of traits was then used to derive a study-wide data-reduced Bonferroni p-value threshold of 0.05/33 or 1.515×10^-3^.

### Genetic data

Genotype data from a total of 2,293 FGFP volunteers were included in the study and a complete description of genotyping and quality control is described in detail in Hughes *et al*. [60]. In brief, 2,646 individuals were genotyped on Illumina Human Core Exome array (v1.0 and v1.1), processed on GenomeStudio v.2.0.4 and quality controlled to remove unmapped variants, duplicated sites, those with missingness greater than 5%, and Hardy–Weinberg equilibrium deviation P<1×10^−5^. Quality control was then performed on samples to remove individuals with array failures, variant missingness greater than 5%, excess heterozygosity and cryptic relatedness. Genotype data was then merged with all Phase 3 data of the 1000 Genomes data set to identify individuals of European genetic ancestry using principal component analysis. Following these quality control steps 2,293 individuals and 509,886 variants remained. Individuals were then imputed to a version of the Haplotype Reference Consortium that included the UK10K and all individuals from Phase 3 of the 1000 Genomes Project.

### MR assumptions

With all MR analyses there are three core assumptions (**Figure 1**). First, the relevance assumption dictates that the instrument(s) must be robustly associated with the exposure. Second, the independence assumption states that there must be no confounders of the instrument(s) and outcome. Third, the exclusion restriction assumption states that the instrument is only related to the outcome via the exposure, that is, there is no horizontal pleiotropy or path from the instrument to the outcome except via the exposure. We have followed STROBE-MR reporting guidelines, and the reporting checklist can be found among online materials [49].

### Construction of a BMI polygenic score

The instrumental variable for BMI used in this study is a polygenic score (PGS). We are explicitly not referring to the PGS as a risk score as BMI itself is not a disease. The PGS was derived from a meta-analysis of genome-wide association studies (GWASs) that was conducted in a sex-combined and (self-reported) sex-specific manner [68]. In total, three PGS instrumental variables were constructed: one for a general European population, one for European females and one for European males. Compliant with the first MR assumption, we used index variants from Pulit *et al.* (taken from Pulit *et al.* **Supplementary Table 1**), which were those defined by the authors as being associated with BMI via conditional and joint association analysis at P<5×10^−9^ [68]. For all index single nucleotide polymorphisms (SNPs) identified by Pulit *et al.* (combined = 670, females = 281, males = 221 SNPs), we extracted data from the FGFP genotype data set in dosage format. After filtering SNPs for presence in FGFP, allele matching, effect allele frequency matching (+/- 10%), imputation quality (info ≥ 0.3) and Hardy-Weinberg Equilibrium testing in aggregate (P<1×10^-8^), a total of 669 (combined), 280 (female), and 221 (male) SNPs remained to construct the three PGSs. The combined, female, and male PGSs were then constructed by weighting each effect allele by the (respective combined, female, and male) effect estimates provided by Pulit *et al.* and summing across all weighted effect alleles carried by an individual. The processing of genotype dosage data into a PGS can be replicated using the function grs_prepare_dosage_data() available in the study’s GitHub repository[78].

### Observational and Mendelian randomization analysis

After combining genotype data (PGS), microbiome traits, anthropometric and clinical variables the study data set consisted of 2,257 individuals (**Supplementary Figure 5**). However, after accounting for missingness in covariables used in association analyses, the maximum sample size in any single linear model was 2,225. GLMs were used to derive conventional observational effect estimates for BMI on microbiota traits. The glm() function from the R stats package was used, BMI was defined as the exposure of interest, and the covariates sex and age were included in all models. When PA or Entero MTs were defined as outcomes a “binomial” family with a “logit” identity was used to model a binomial distribution and perform a logistic regression in the GLM. When Div, AB, or RA MTs was defined as outcomes, a “gaussian” family with an “identity” link was used to model a normal distribution. In addition, all Div, AB, and RA MTs were rank-based inverse normal transformed prior to running the GLM. After modeling, a Breusch-Pagan test of homoskedasticity was performed using the bptest() function from the ‘lmtest’ R package [80]. A Wald test was performed using the waldtest() function from the ‘lmtest’ R package to extract an F-statistic and P-value for the exposure [80]. A Type II ANOVA was run using the function Anova() from the ‘car’ R package from which the sum of squares (deviances) were extracted [81]. Those sums of squares where then used to estimate an eta-squared (η^2^) statistics for the model and for the exposure. η^2^ provides a measure of the proportion of variance in the outcome explained by each covariate in the model. When testing for a difference between male and female effect estimates we performed a z-test using the bespoke function ztest() found in the study’s GitHub repository [78].

One-sample MR analyses were then performed to estimate the causal effect of BMI on each MT (**Figure 1**). To do so, stage one of the two-stage least squares (MR) model was run by defining BMI as the outcome, the PGS as the exposure and including sex and age as covariates – again using the glm() function. Stage two of the two-stage least squares (MR) model was then estimated using the ivglm() function of the ‘ivtools’ R package using the estimation method “ts” or two-stage which derives the causal effect estimates and standard errors [82]. The ivglm() function takes as input the stage one model and the observational model described above. A Breusch-Pagan test, Wald Test, Wu-Hausman endogeneity test, Type II ANOVA and η^2^ statistics were then run and derived on each MR model. All observational and MR analyses described in this section were run with the bespoke wrapper function ivtoolsfit() that can be found in the study’s GitHub repository [78].

MR analyses were conducted in a sex-combined and sex-specific frameworks. For the sex-specific analyses we used both (1) the general population (sex-combined) PGS as the instrument, and (2) the sex-specific PGSs. The use of the overall PGS in sex-specific frameworks provides, as we interpret it, estimates for the effect of life-course elevated risk of higher BMI, as modeled in an average adult human, on MT variation in the individual sexes. Sex-specific PGSs were used to capture sexual dimorphic differences in adiposity distribution, reflected by BMI [83], that a general population-based PGS averages out.

For sensitivity analyses, we repeated the observational and MR analyses as described above but include 16S total read count (the number of sequencing reads prior to being rarefied), Bristol stool score, J01 antibiotic use (yes = 1, no = 0), smoking status (never, ever, current), and household monthly income as additional model covariates (**Table 1**). Total read counts, Bristol stool score, and antibiotic use have known large effects on our outcome and are being included as precision variables to reduce technical or measurement error in both observational and MR analyses and to theoretically assess possible confounding in the observational analyses. Smoking status and household income, in contrast, are known to associate with our exposure and may be possible confounders of our exposure-outcome relationship in observational analyses [18].

**Table 1.** FGFP Population description. **Table 1**: Population descriptive summary statistics for the FGFP cohort as a whole (overall), and by sex (females and males). Reported are mean and (95% quantile intervales) for age, height, weight, BMI. Also, the number (percent) of individuals who were never, ever, and current smokers, and the number (percent) of individuals across monthly household income categories are presented.

| Characteristic | N | Overall<br>N = 2,228<sup>1</sup> | female<br>N = 1,337<sup>1</sup> | male<br>N = 891<sup>1</sup> |
| --- | --- | --- | --- | --- |
| **age** | 2,228 | 53 (25, 73) | 51 (24, 72) | 55 (27, 75) |
| **height** | 2,228 | 170 (154, 188) | 166 (153, 178) | 177 (165, 190) |
| **weight** | 2,228 | 72 (50, 106) | 66 (48, 95) | 81 (60, 112) |
| **BMI** | 2,228 | 24.8 (18.4, 34.9) | 24.2 (17.9, 35.0) | 25.8 (19.3, 34.8) |
| **smoking** | 1,991 |  |  |  |
| never |  | 1,024 (51%) | 705 (58%) | 319 (41%) |
| ever |  | 798 (40%) | 412 (34%) | 386 (49%) |
| current |  | 169 (8.5%) | 93 (7.7%) | 76 (9.7%) |
| **household income** | 1,940 |  |  |  |
| I_dont_know |  | 49 (2.5%) | 38 (3.2%) | 11 (1.4%) |
| rather_not_answer |  | 218 (11%) | 152 (13%) | 66 (8.7%) |
| less_than_750 |  | 8 (0.4%) | 6 (0.5%) | 2 (0.3%) |
| 1000_to_1500 |  | 118 (6.1%) | 79 (6.7%) | 39 (5.1%) |
| 1500_to_2000 |  | 194 (10%) | 117 (9.9%) | 77 (10%) |
| 2000_to_2500 |  | 289 (15%) | 172 (15%) | 117 (15%) |
| 2500_to_3000 |  | 275 (14%) | 149 (13%) | 126 (17%) |
| 3000_to_3500 |  | 274 (14%) | 176 (15%) | 98 (13%) |
| more_than_3500 |  | 515 (27%) | 288 (24%) | 227 (30%) |

<sup>1</sup> Mean (Q2.5, Q97.5); n (%)

Given that the exposure trait – in its published GWAS - and all outcome abundance traits were rank-based inverse normal transformed prior to running linear models all effect estimates for AB, RA and diversity traits represent a rank normal standard deviation unit of change for each rank normal standard deviation unit increase in BMI. Effect estimates for binary PA traits are presented in log-odds, such that they represent the log-odds of presence (or being assigned to Enterotype class ‘X’; coded as 1=Enterotype ‘X’, 0=other) for each rank normal standard deviation unit increase in BMI. Throughout the manuscript, references to increases in BMI or MTs reflect cross-sectional differences, not longitudinal changes within individuals over time.

### Code and data availability

All statistical analysis were conducted in the R language (v 4.0.2, Taking Off Again) and all bespoke functions and analytical code for the study can be found in the GitHub repository https://github.com/hughesevoanth/FGFP_BMI_MR [78,84]. FGFP genotype data and host metadata are not open access but are available in accordance and in consent with ethical permission through managed access subject to a data use agreement with the FGFP and organized via principal investigator J.R. Raw 16S data are available at the European Genome/Phenome Archive under accession no. EGAS00001004420.

## Results

### Population description

The sample population, with data availability, consisted of 2,228 individuals, of which 1,337 were female and 891 were male (**Table 1**). The sample population mean BMI (exposure) was 24.8 kg/m^2^ (95% quantile interval (QI): 18.4-34.9), with males (25.8 kg/m^2^) having a higher average BMI than females (24.42 kg/m^2^, T-test *p* = 2.8×10^-19^). Fifty-eight percent of the sample population had a “healthy” BMI (<25 kg/m^2^; 47% for males, 65% for females), 30% had “overweight” (BMI ≥25 & <30 kg/m^2^; 39% for males, 24% for females) and 12% had “obesity” (BMI >30 kg/m^2^; 14% for males, 11% for females).

A total of 368 primary MTs were included in observational and MR association analyses. This includes 72 abundance (AB) MTs, 188 ratio (RA) MTs, and 108 binary (PA) MTs. The AB traits include two β-diversity, three alpha-diversity and 67 abundances (19 of which are trunc_AB). The PA traits included 104 PA traits and four binary enterotype classes. For simplicity, in text and figures, we will refer to all MT ratio outcomes as ‘RA’, all other MTs that were modeled with a gaussian distribution as ‘AB’ and all MT outcomes modeled with a binomial distribution as ‘PA’, unless specified otherwise. Summary statistics and trait type description for all MTs, including presence/absence counts, proportion of zero observations in a trait, minimum, mean, maximum, kurtosis, and Shapiro-Wilks W-statistic are available in **Supplementary Table 1**.

### Observational associations

In observational analyses, 141 MTs were associated with BMI, with 74 positive and 67 negative associations, across 39 PA, 78 RA, and 24 AB traits (including all five diversity metrics and three enterotypes; **Figure 2A, 3, and Supplementary Table 3**). We found that increases in BMI were associated with decreased diversity for Shannon (β = -0.022, se = 0.005, *p* = 1.23×10^-5^), number of genera (β = -0.021, se = 0.005, *p* = 2.58×10^-5^) and Chao1 (β = -0.021, se = 0.005, *p* = 2.92×10^-5^) diversity metrics (**Figure 2A, 3, and 4C**), where β reflects rank-normalized standard deviation units of change per 1 kg/m² increase in BMI. In addition, BMI was associated with β-diversity showing an inverse association with MDS1 (multiple dimension scaling axis 1; β = -0.027, se = 0.005, *p* = 1.00×10^-7^) and a positive association with MDS2 (β = 0.024, se = 0.005, *p* = 9.00×10^-7^; **Figure 4B and C**). Connected to this, BMI was associated with three of the enterotype classes; assignment to enterotypes Bacteroides 2 (β = 0.045, se = 0.013, *p* = 3.43×10^-4^) and Prevotella (β = 0.048, se = 0.013, *p* = 2.00×10^-4^) positively associated with BMI while assignment to enterotype Ruminococcus (β = -0.061, se = 0.012, *p* = 5.26×10^-7^) was inversely associated with BMI.

**Figure 2.**
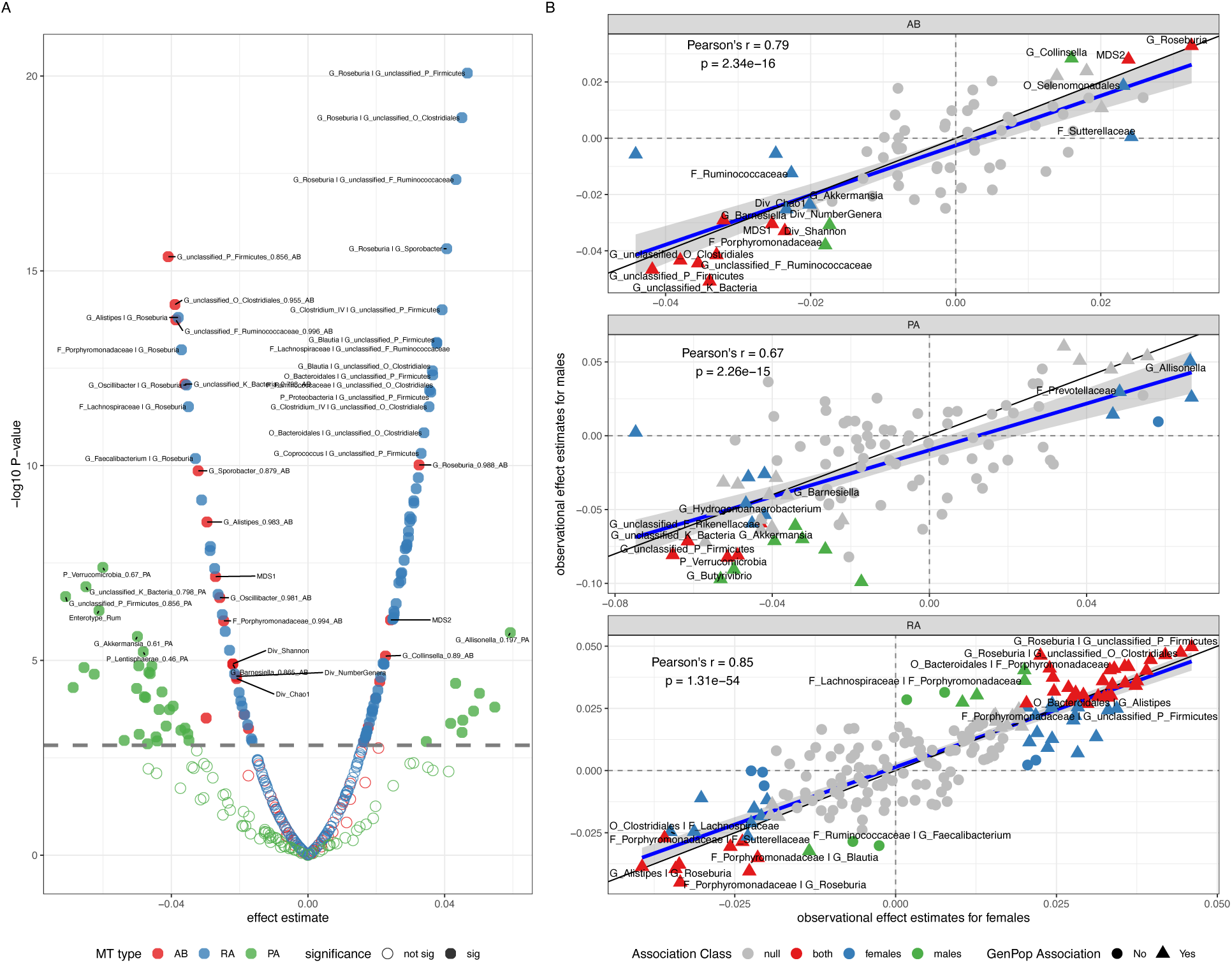
Observational effect estimates. (A) A volcano plot of the general population effect estimates for the association of BMI and microbiota traits. The x-axis denotes the effect estimates, in rank inverse normalized standard deviation units of change per unit increase in BMI (kg/m^2^) for AB (abundance; in red) and RA (ratio; in blue) traits, or in odds of presence per unit increase in BMI (kg/m^2^) for PA (presence | absence; in green) traits. The y-axis denotes the negative log10 of the association p-value. The horizontal dashed line denotes the study’s alpha threshold for declaring an association. (B) Scatter plots of the correlation between female-derived (x-axis) and male-derived (y-axis) BMI observational effect estimates for AB (top), PA (middle), and RA (bottom) microbiota traits. Microbiota traits that were associated with BMI in the general population are illustrated as triangles, and those that were not, are illustrated as circles. Those microbiota traits that associated with BMI in both female and male specific frameworks are colored red, those associated with BMI in only females are blue, and only males are green. The microbiota traits that are colored grey were not associated with BMI in either the female or male specific analyses. Pearson’s correlation coefficients and p-values, for the correlation between female and male observational effect estimates, are denoted in the top left-hand corner of each plot.

**Figure 3:**
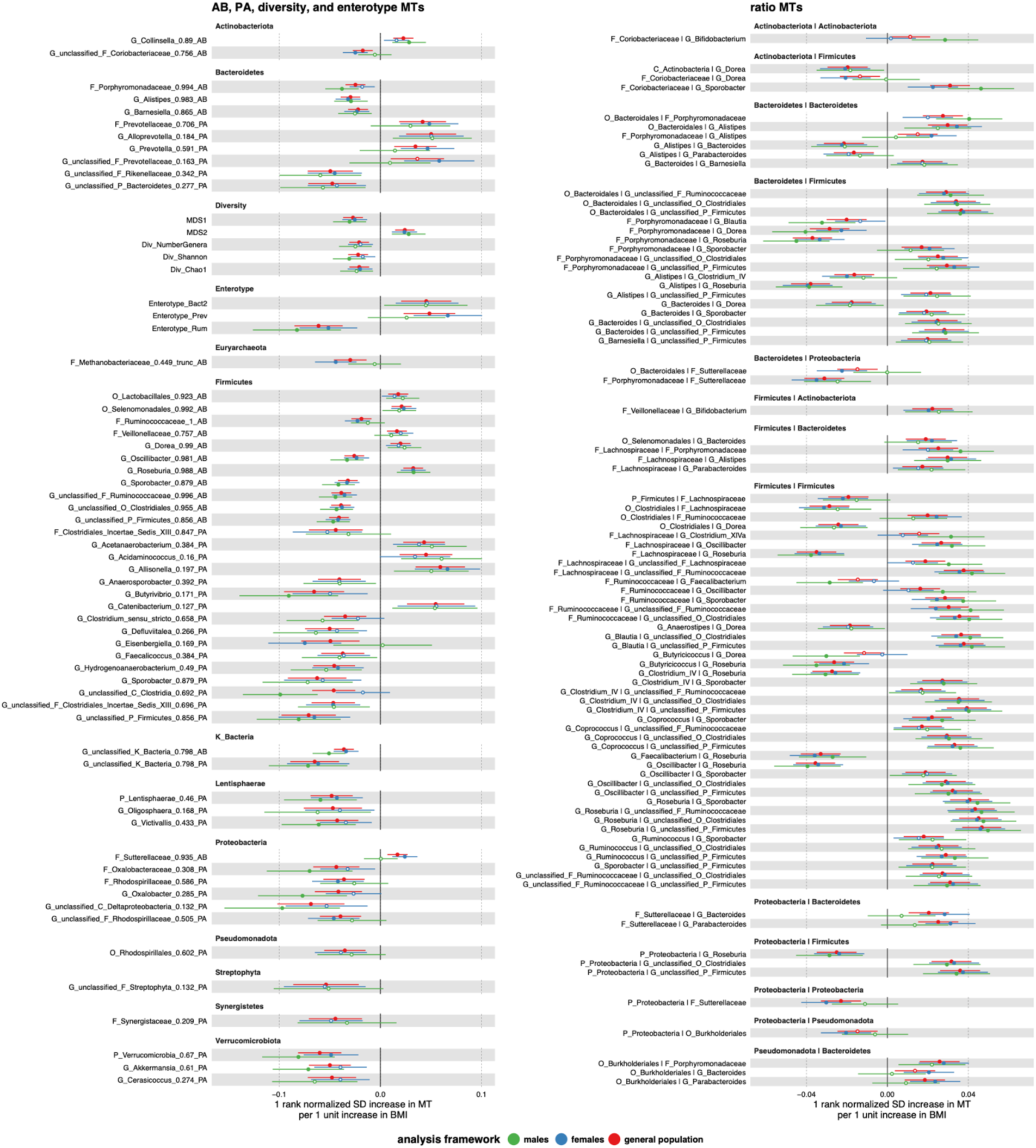
Forest plot of observational associations for AB and PA MTs. Observational effect estimates for BMI on MTs are illustrated as dots along with their 95% confidence intervals (bars). Effect estimates for the general population (red), females (blue), and males (green) are illustrated for each MT. Point estimates that are solid in color surpassed our multiple testing burden to declare an association (P>0.05/33), while those with a white dot did not. MTs are organized by their phylum or trait type.

**Figure 4:**
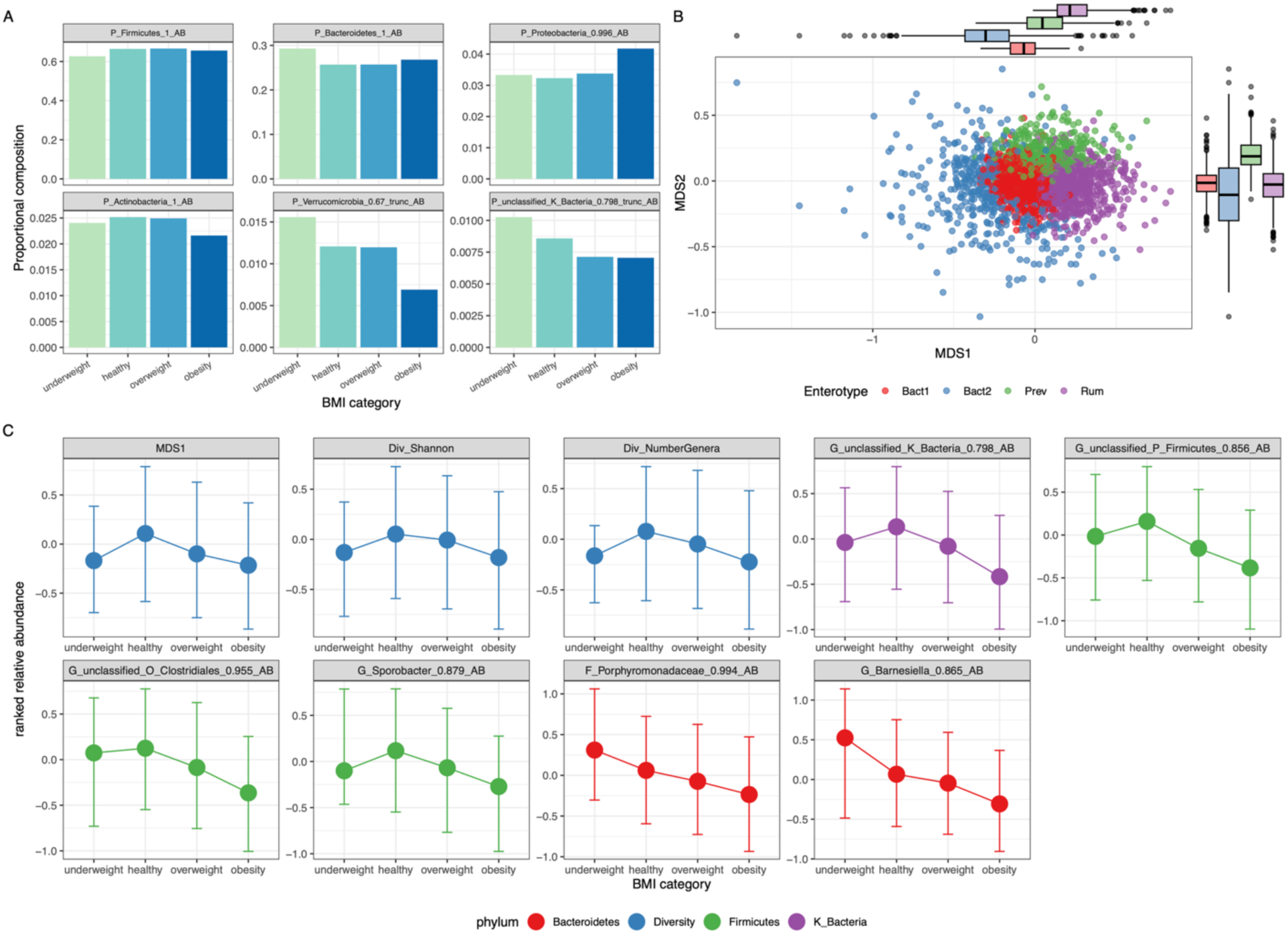
Phyla abundance, enterotype and MT change with BMI category. (A) A bar plot of mean relative abundance for 6 common phyla for four BMI categories; underweight (BMI <= 18), healthy (18 < BMI <= 25), overweight (25 < BMI <= 30), and obesity (BMI > 30). (B) A multidimensional scaling plot of the FGFP data set. The x and y-axis are the Beta-diversity metrics MDS1 and MDS2, respectively, used in the study. Individual samples are color coded by their assignment to one of the four enterotype classes (Bact1, Bact2, Prev, and Rum), the distributions of which are represented by the box-plots on the x and y axis of the scatter plot. (C) Median rank inverse normal transformed relative abundance point estimates (circles) and the interquartile range (bars) for each microbiota trait by BMI category. Microbiota traits, causally associated with BMI (MR) and chosen as exemplars, are indicated by name in the title of each individual plot and color coded by their associated phylum, kingdom, or trait type as indicated by the key at the bottom of the figure.

At the genera level, seven AB MTs associated with BMI. They were *Collinsella* of phylum Actinobacteriota, *Alistipes* and *Barnesiella* of phylum Bacteroidetes, and *Dorea*, *Oscillibacter*, *Roseburia* and *Sporobacter* of phylum Firmicutes. Three genera, namely *Dorea* (β = 0.020, se = 0.005, *p* = 9.40×10^-5^) and *Roseburia* (β = 0.033, se = 0.005, *p* = 9.64×10^-11^) of phylum Firmicutes, and *Collinsella* (β = 0.023, se = 0.005, *p* = 7.70×10^-6^) of phylum Actinobacteriota, were each positively associated with BMI (**Figure 2 and 3**). Here, the butyrate producing genus *Roseburia* had the most pronounced positive association with BMI. Conversely, the genera-level AB MTs *Alistipes* (β = -0.030, se = 0.005, *p* = 2.80×10^-9^) and *Barnesiella* (β = -0.022, se = 0.005, *p* = 1.25×10^-5^) of phylum Bacteroidetes, and *Oscillibacter* (β = -0.026, se = 0.005, *p* = 2.45×10^-7^) and *Sporobacter* (β = -0.032, se = 0.005, *p* = 1.36×10^-10^) of phylum Firmicutes were inversely associated with BMI. Here, *Sporobacter* had the strongest inverse association. Interestingly, two genera of phylum Firmicutes (*Dorea* and *Roseburia*) were positively associated with BMI while two others (*Oscillibacter* and *Sporobacter*) were inversely associated with BMI. Numerous unclassified AB MTs also exhibited strong inverse associations, however it remains difficult to make any strong biological inferences about these observations as these unclassified collections of 16S rRNA reads cannot be assigned beyond the taxonomic level identified. For example, the AB MT with the strongest association with BMI was G_unclassified_P_Firmicutes (β = -0.041, se = 0.005, *p* = 4.34×10^-16^). This MT is composed of all 16S rRNA reads that could be assigned to phylum Firmicutes, but to no lower phylogenetic level. As a product these reads may belong to one or many different genera.

There was very little observed evidence of an association between the ratio of phylum Firmicutes and phylum Bacteroidetes (β = -0.004, se = 0.005, *p* = 0.39; **Figure 4A**) and BMI. Nor was there an association with either of the AB MTs of these phyla (P_Bacteroidetes_1_AB: β = 0.005, se = 0.005, p = 0.31; P_Firmicutes_1_AB: β = 0.0004, se = 0.005, *p* = 0.93). There were however, 78 associations between RA MTs and BMI. This includes 21 Firmicutes-to-Bacteroidetes RA MTs made up of AB MTs from lower taxonomic units. For example, the strongest observed RA MT derived from lower order taxa of phyla Bacteroidetes and Firmicutes was G_Alistipes | G_Roseburia_RA (β = -0.038, se = 0.005, *p* = 1.58×10^-14^). In addition, a total of ten Firmicutes-to-Bacteroidetes RA MTs increased and 11 decreased with increases in BMI. These observational RA results in combination with the AB and PA results for these phyla suggest that there is not a simple, singular directional relationship between Firmicutes, Bacteroidetes and BMI - given that we observe both negative and positive associations across phyla and trait types (AB, PA, and RA, **Figure 3)**.

In sex-specific analyses, we observed effect estimates (**Supplementary Table 3**) to be, in general, similar between males and females (Pearson’s r = 0.77 across all primary traits, **Figure 2B**), supporting broad directional consistency among estimates. Though there are some specific differences. In total, we observed 99 MTs to be associated with BMI in females and 72 MTs to be associated with BMI in males – 53 of which are shared between the sexes (**Figure 2B**). Ninety-three of the 99 female-associated MTs, 68 of the 72 male-associated MTs, and all 53 of the shared MTs, were associated with BMI in the aggregated analysis reported above. The 53 shared MTs include the β-diversity metric MDS1 and MDS2, Enterotype Ruminococcus, the genera AB MTs *Sporobacter*, *Roseburia*, *Alistipes*, and *Oscillibacter*, and the phylum PA MTs Verrucomicrobia and Lentisphaerae. All of which, except for *Roseburia* and MDS2, are negative associations (**Figure 2B and 3**). The four male associations not observed in the combined analysis were all ratio traits, that include G_Butyricicoccus | G_Dorea_RA (male β = -0.030, se = 0.008, *p* = 3.94×10^-4^; female β = -0.002, se = 0.006, *p* = 0.68). The six female associations not observed in the combined analysis included five RA and 1 PA MT, including O_Bacteroidales|F_Sutterellaceae_RA (female β = -0.022, se = 0.006, *p* = 4.25×10^-4^, male β = - 0.000, se = 0.009, *p* = 0.99).

### Mendelian randomization

Three BMI polygenic scores (PGSs), one overall, a second for females and a third for males were constructed from 669, 280, and 221 genetic variants previously associated (*p* < 5×10^−9^) with BMI, respectively (see **Materials and Methods**) [68]. The overall PGS explains 4.65% of the variation in BMI in the general population, 3.90% of the variation in BMI among females, and 6.56% of the variation in BMI among males – estimates were derived from univariable models. The female-specific PGS explains 2.63% of the variation in BMI among females and the male-specific PGS explains 3.88% of the variation in BMI among males – in univariable models. Each PGS was used as an instrumental variable for BMI in MR analysis providing estimates of the effect of BMI on MT outcomes.

Overall, (primary) MR estimates (n = 368) were strongly correlated with observational estimates, with a Pearson’s r of 0.65 (*p* = 5.29×10^-10^) among AB MTs, a Pearson’s r of 0.79 among RA MTs (*p* = 6.34×10^-42^), and a Pearson’s r of 0.55 among PA MTs (*p* = 6.78×10^-10^, **Figure 5B**). As expected, MR (life-course) effect estimates were generally larger than observational estimates (Wilcoxon test: mean absolute difference = 0.038, 95% QI 0.001-0.123, *p* = 4.65×10^-62^). Results from the Wu–Hausman endogeneity test indicated that most models were consistent with BMI being exogenous. However, nine models exhibited evidence of endogeneity (p < 0.05/33; **Supplementary Table 3**), which may indicate confounding in the observational estimates.

**Figure 5:**
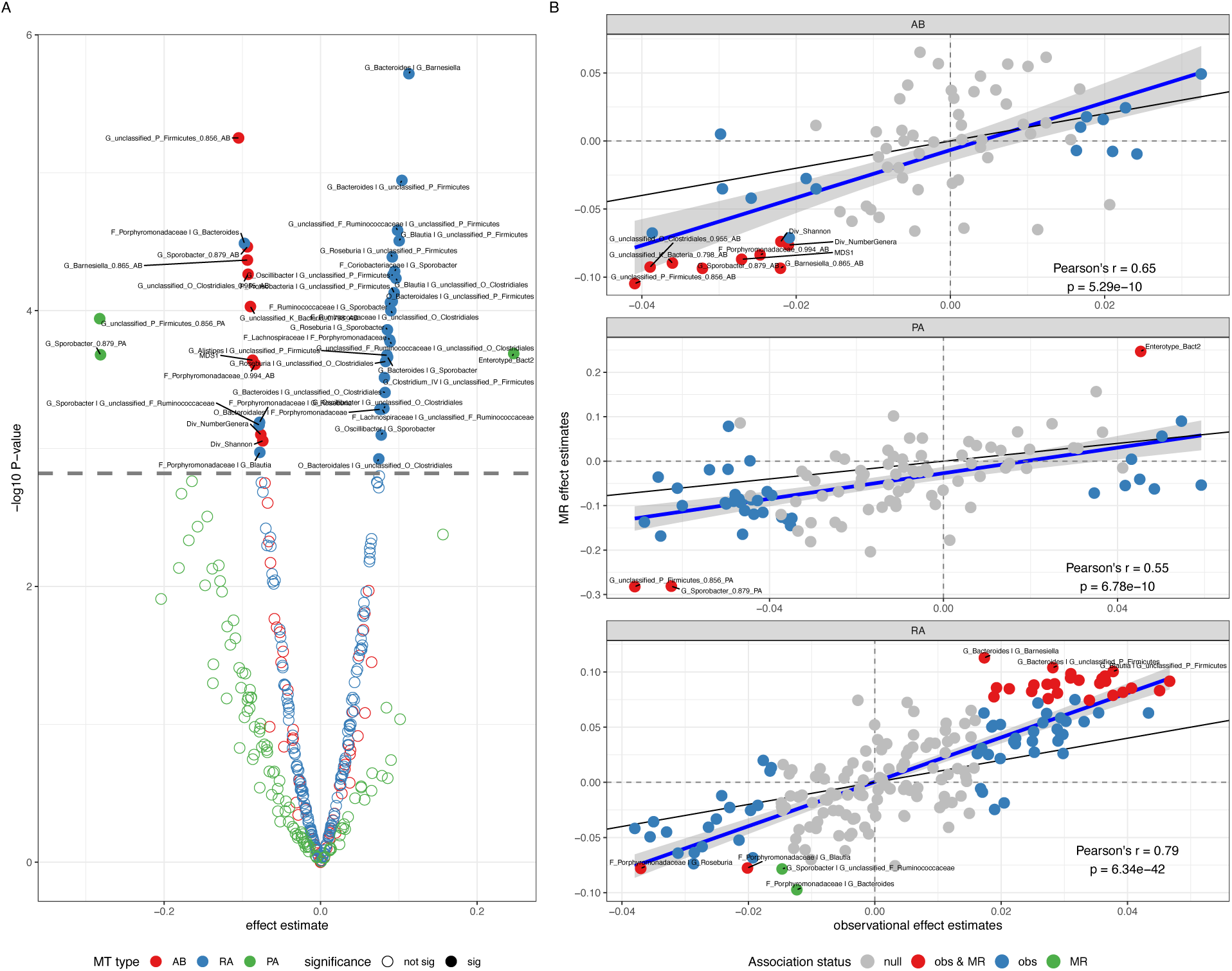
MR effect estimates: (A) A volcano plot of the general population MR effect estimates for BMI on microbiota. The x-axis denotes the effect estimates, in rank inverse normalized standard deviation units of change per unit increase in BMI (kg/m^2^) for AB (abundance; in red) and RA (ratio; in blue) traits, or in odds of presence per unit increase in BMI (kg/m^2^) for PA (presence | absence; in green) traits. The y-axis denotes the negative log10 of the MR p-value. The horizontal dashed line denotes the study’s alpha threshold for declaring a causal effect. (B) Scatter plots of the correlation between observational (x-axis) and MR (y-axis) effect estimates for BMI on AB (top), PA (middle), and RA (bottom) microbiota traits. Microbiota traits that associated with BMI in both observational and MR frameworks are colored red, those associated with BMI in only observational analyses are blue, and only in MR analyses are green. The microbiota traits that are colored grey were not associated with BMI in either framework. Pearson’s correlation coefficients and p-values, for the correlation between observational and MR effect estimates, are denoted in the bottom right-hand corner of each plot.

There was evidence of a causal relationship between BMI and 41 MTs. This included three PA (including one enterotype), nine AB (including three diversity metrics) and 29 RA MTs (**Table 2**, **Figure 5A and 6**, **Supplementary Table 3**). Among the 41 MTs with evidence of a causal relationship, six exhibited evidence of endogeneity in the Wu–Hausman test, indicating that observational and MR estimates differed for these traits. Although observational and MR effect estimates were all directionally consistent.

**Figure 6:**
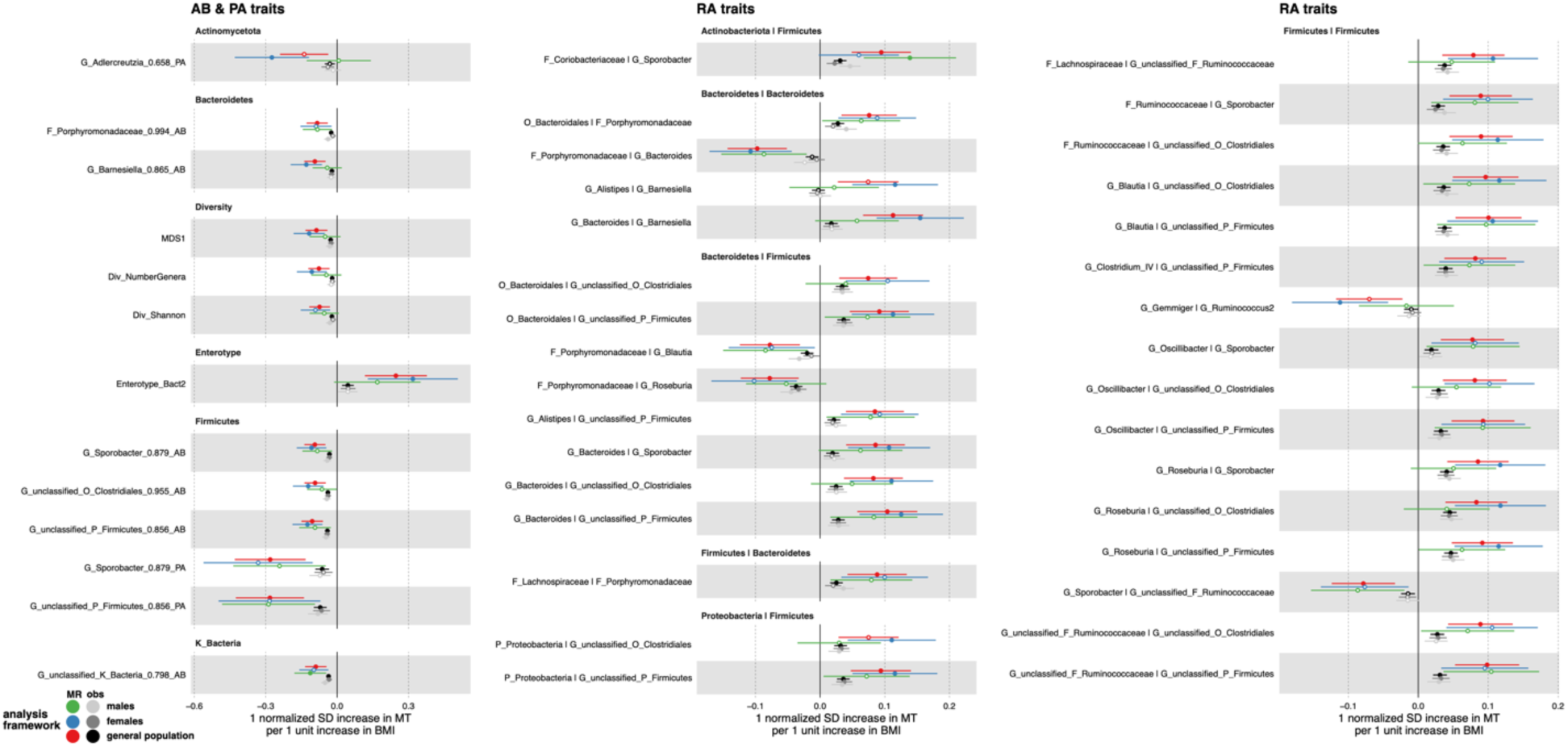
Forest plot of MR effect estimates. MR and observational effect estimates for BMI on MTs are illustrated as the dots along with their 95% confidence intervals. MR effect estimates for the general population (red), females (blue), and males (green) are illustrated for each MT. In addition, observational effect estimates for the general population (black), females (dark grey), and males (light grey) are also presented for each MT. Point estimates that are solid in color surpassed our multiple testing burden to declare an association (P>0.05/33), while those with a white dot did not. MTs are organized by their phylum or trait type.

**Table 2:**
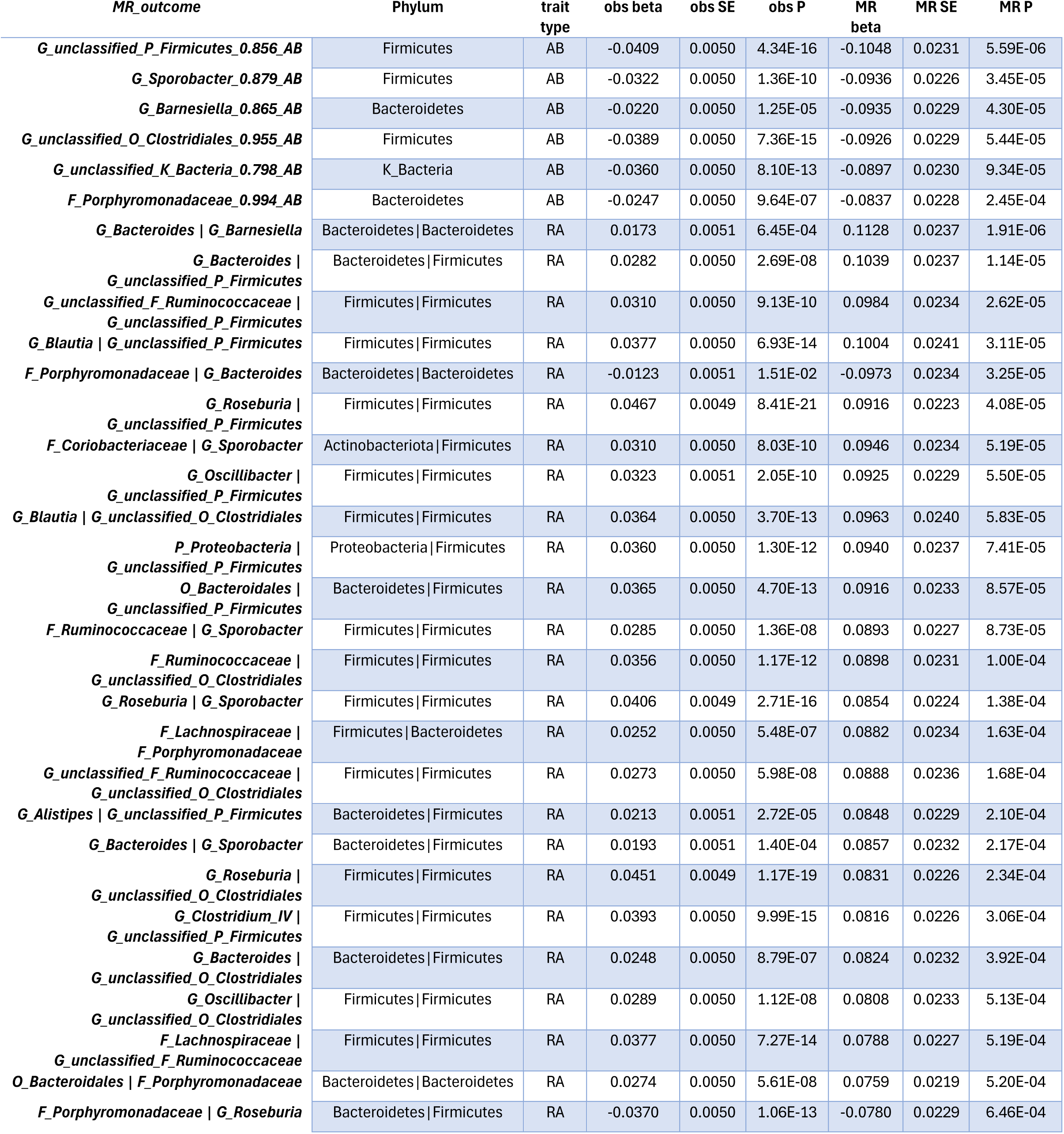
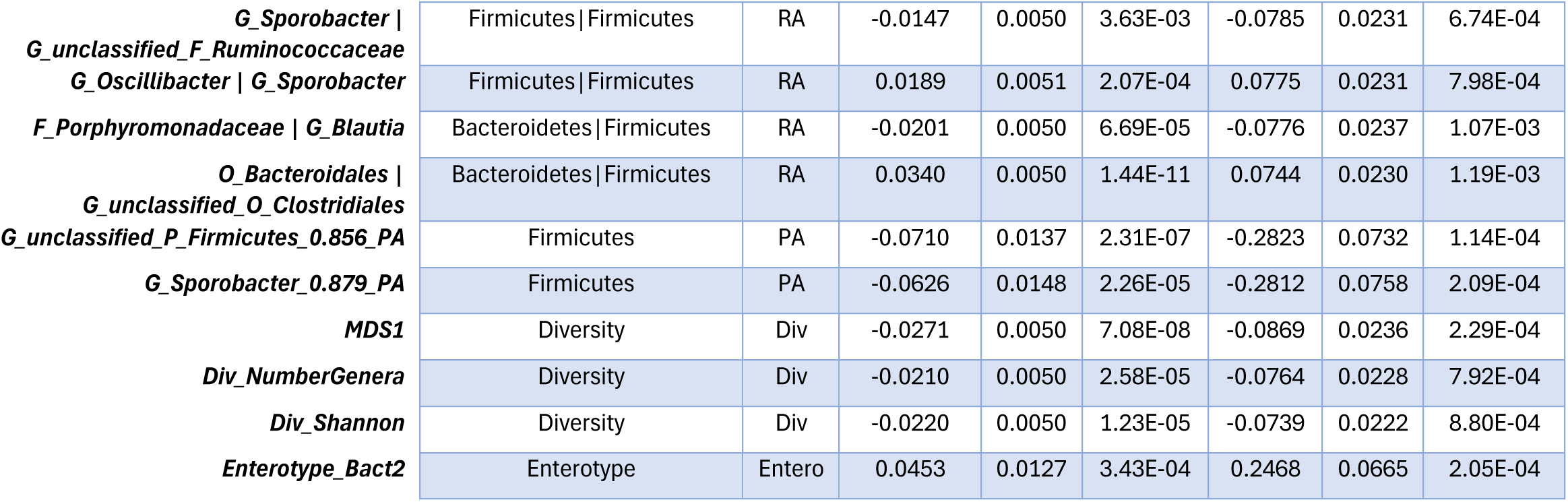
Microbiota traits associated with BMI. **Table 2, Microbiota traits influenced by BMI:** The 28 microbiota traits with evidence suggesting that BMI influences their variation as determined by a one-sample MR framework. Presented are the MT, the phylum(s) from which the MTs are derived or their specific class, as well as their trait type (AB=abundance, RA=ratio, PA=presence|absence, Div=diversity, Entero=enterotype). In addition, the effect estimate (beta), standard error (se), and p-value (P) are provided for the observational (Obs) and Mendelian Randomization (MR) frameworks.

Alpha-diversity metrics, including the number of genera (β = -0.076, se = 0.023, *p* = 7.92 x10^-4^), and Shannon diversity (β = -0.074, se = 0.022, *p* = 8.80×10^-4^) both decreased with increased BMI. This is consistent with higher BMI causally decreasing gut microbiome diversity. In addition, the abundance of family *Porphyromonadaceae* (β = -0.084, se = 0.023, *p* = 2.45×10^-4^), genus *Barnesiella* (β = -0.094, se = 0.023, *p* = 4.30×10^-5^) and genus *Sporobacter* (β = -0.094, se = 0.023, *p* = 3.45×10^-5^) inversely associated with BMI, as did the abundance of unclassified kingdom Bacteria (β = -0.090, se = 0.023, *p* =9.34×10^-5^), unclassified phylum Firmicutes (β = -0.105, se = 0.023, *p* = 5.59×10^-6^) and unclassified order Clostridiales (β = -0.093, se = 0.023, *p* =5.44×10^-5^) of phylum Firmicutes. Further, β-diversity as measured by MDS1 (β = -0.087, se = 0.024, *p* = 2.29×10^-4^) decreased and assignment to enterotype Bacteroides2 increased with higher BMI (β = 0.25, se = 0.066, *p* = 2.05×10^-4^). Genus *Barnesiella* (Wu-Housman test *p* = 1.28×10^-3^) and enterotype Bacteroides2 (*p* = 4.55×10^-4^) were two traits that exhibited evidence of endogeneity.

The 29 RA MTs with evidence of a BMI causal effect mapped to 17 taxa across four phyla. Two phyla, Firmicutes and Bacteroidetes, and more specifically two orders within these phyla, Bacteriodales (n=12) and Clostridiales (n=22), respectively, make up 14 of the 17 involved taxa (**Figure 6 and 7**). These RA MTs suggest correlated changes either within, between or involving these two phyla (**Figure 6 and 7**). Eight genera were involved in BMI associated RA MTs including: Bacteroides (n=5), Alistipes (n=1) and Barnesiella (n=1) of phylum Bacteroidetes, and Sporobacter (n=7), Roseburia (n=4), Blautia (n=3), Oscillibacter (n=3) and Clostridium IV (n=1) of phylum Firmicutes. In general, these RA MT observations suggest a dynamic trade-off system within the microbial community, even within the same phylum. For example, we observed that the ratio of order Bacteroidales to family *Porphyromonadaceae* (of order Bacteroidales; O_Bacteroidales | F_Porphyromonadaceae) increases with increased BMI. This suggests that as the abundance for family *Porphyromonadaceae* decreases there is corresponding increase in reads to other family taxa within the same order, such as those from genus *Bacteroides* of family Bacteroidaceae (F_Porphyromonadaceae | G_Bacteroides; **Figure 6 and 7**).

**Figure 7.**
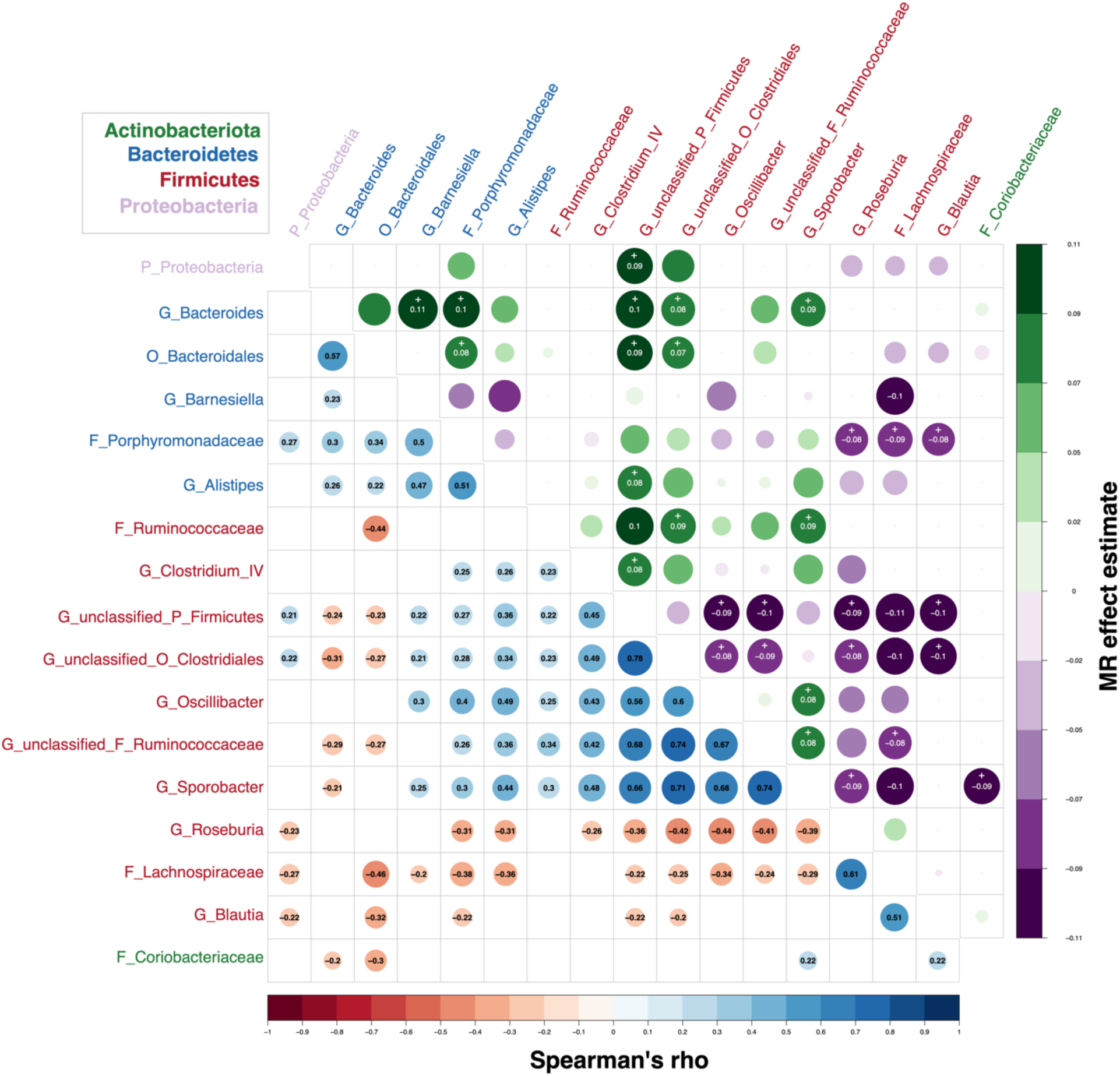
Correlation among MTs included in RA MTs and influenced by BMI. Twenty-nine RA MTs exhibited evidence of BMI influencing their variation in a one-sample MR framework. Seventeen AB MTs were involved in these 29 associated RA traits. Those 17 AB MTs are included in this plot. The lower triangle presents the Spearman’s rho between all pairs of these AB MTs. Those lower-triangle cell with a numerical value in them, indicate pairs of AB MTs that met our criteria to build a RA MT. The upper triangle presents the MR effect estimates for BMI on the RA MTs. The ratios, as illustrated in this plot, have the AB MT on the x-axis in the numerator and the MT on the y-axis in the denominator of the RA. Cells in the upper triangle with a numerical value indicate those that surpass our multiple test correction threshold to declare an association with BMI in the MR analysis. In addition, upper-triangle cells with a ‘+’ indicate those 29 associated RA MTs that were included in the “primary traits” list and were not considered “redundant” MTs strongly correlated with another MT in that list. As such the RA G_Roseburia | G_unclassified_O_Clostridiales has an MR effect estimate of 0.08, indicating BMI has a causal effect on that MT and it also has a ‘+’ in its upper triangle indicating this RA was included in study’s “primary traits”. In addition, the cell in the lower triangle for these two AB MTs (G_Roseburia and G_unclassified_O_Clostridiales) indicate a Spearman’s rho of -0.42. Taxa names along the x and y axis are color coded by their phylum as indicated by the key in the top left corner.

### Sex-specific Mendelian randomization

We repeated the MR analyses using the overall PGS (PGS_genpop_) within a sex-specific framework (**Supplementary Table 3**). This approach estimates the causal effect of higher BMI, as modeled in an average adult, on MT variation specific to each sex. Overall, female-specific MR estimates were broadly consistent in magnitude and direction with the general population estimates (Pearson’s r = 0.92, **Supplementary Figure 6**). Twenty-five MTs associated with BMI in females: six AB traits (including two Div measures), seventeen RA traits, and two PA traits (including one Entero). Four associations in females were novel and not observed in the general population analysis, though they were directionally consistent with the general population estimates (**Figure 6**). These novel associations included a PA MT G_Aldercreuzia_0.658_PA (β = -0.274, se = 0.079, p = 5.69×10^-4^), and three RA MTs: G_Alistipes | G_Barnesiella (β = 0.116, se = 0.034, p = 6.04×10^-4^), G_Gemmiger | G_Ruminococcus2 (β = -0.112, se = 0.035, p = 1.42×10^-3^), and P_Proteobacteria | G_unclassified_O_Clostridiales (β = 0.111, se = 0.035, p = 1.44×10^-3^). The first two RA MTs again illustrate inverse abundance relationships or trade-off dynamics among genera within the same phylum. Notably, very little evidence of an effect was found in males for any of these four female specific associations, as each had near zero point estimates. Most striking was the difference in the size of the effect estimate for MT G_Aldercreuzia_0.658_PA between the sexes, which was β = 0.007 (se = 0.069, p = 0.92) for males (**Figure 6** and **Supplementary Figure 7**).

Male-specific MR estimates (using PGS_genpop_) were also broadly consistent with the general population estimate (Pearson’s r = 0.82) (**Supplementary Figure 6**). Two MTs were associated with BMI in males: G_unclassified_K_Bacteria_0.798_AB (β = -0.113, se = 0.034, p = 9.50×10^-4^) and F_Coriobacteriaceae | G_Sporobacter (β = 0.139, se = 0.036, p = 1.37×10^-4^). Both MTs were identified in the general population analysis but not in females, although their effect estimates were directionally consistent between sexes. Effect estimates between males and females correlated moderately well (Pearson’s r = 0.549, p = 2.70×10^-30^), yet estimates were on average larger in females than males (absolute mean difference = 0.054, 95% quantile interval (QI) = 0.004-0.172, Wilcoxon test p = 3.33×10^-111^).

We repeated sex-specific MR analyses using sex-specific BMI PGSs as instrumental variables for females and males, respectively. Unlike the previous analyses, these additional MR analyses were conducted because sex-specific PGSs may capture sexual dimorphic differences in adiposity distribution, reflected by BMI [83], that a general population-based PGS could average out. In short, these analyses may better account for sex difference in both BMI and MT variation. Overall, MR estimates obtained using sex-specific PGSs correlated broadly with sex-specific estimates obtained using the general population PGS: Pearson’s r = 0.86 for females, and Pearson’s r = 0.64 for males (**Supplementary Figure 8**). No MTs were associated with BMI in males. However, three MTs were associated with BMI in females: G_unclassified_F_Coriobacteriaceae_0.756_AB (β = -0.152, se = 0.046, p = 8.71×10-4), F_Coriobacteriaceae | G_Ruminococcus2 (β = -0.164, se = 0.049, p = 7.29×10-4), and G_Bacteroides | G_Barnesiella (β = 0.138, se = 0.043, p = 1.28×10-3) (**Supplementary Table 3**). The later association, G_Bacteroides | G_Barnesiella, was also observed in both the general population and female (PGS_genpop_) analyses presented above. However, the first two associations were uniquely identified in this analysis, though their directions of effect remained consistent in both the general population and female (PGS_genpop_) analyses (**Supplementary Figure 7**). In addition, there was little or no evidence of an association between BMI and these two MTs in males (**Supplementary Figure 7**).

### Sensitivity analyses

We repeated both observational and MR analyses by including 5 additional covariates total read count, Bristol stool score, J01 antibiotic use, smoking status, and household income. Each have been previously associated with either our exposure, outcome, or both. As such they are being included as precision variables and to account for possible confounding of the exposure-outcome association in observational analyses. However, we note that individual-level confounding variables could potentially violate the third MR assumption (exclusion restriction) through horizontal pleiotropy, if there existed an association between the genetic variants and these variables. We also note that, if there are unknown population-level confounders that violate the second MR assumption (independence), then the inclusion of these additional individual-level covariates could induce collider bias in MR analyses if there existed an association between these population- and individual-level confounders. Best practice to avoid violation of the independence assumption includes the use of homogenous population samples - in both the exposure GWAS and the one sample MR sample populations - where population confounding due to structure and/or stratification have been minimized. Homogenous sample populations, however, are difficult to achieve [85]. Nevertheless, the inclusion of these sensitivity analyses allow us to gain additional insights from these adjustments. We do not assume that one model is better than another.

In both conventional observational and MR sensitivity analyses (**Supplementary Table 4**), primary MT effect estimates were highly concordant with those from the primary analysis with a Pearson’s correlation coefficient of 0.959 and 0.938, respectively. The Pearson’s r among the 41 general population MR associations was 0.99 and the largest p-value observed in the sensitivity analysis among these 41 MR associations was 0.023. Overall, these results suggests that the inclusion of these variables would not greatly alter our conclusion.

### Untransformed analyses

In addition, for future comparative convenience, all primary association analyses were conducted without rank inverse normal transforming the outcome (**Supplementary Table 5**). However, we warn that many of the MT outcome distributions are far from being appropriate for a parametric model. As a result, special attention should be given to the Shapiro-Wilks W-statistics of the model residuals available in the supplementary table.

## Discussion

Analyses undertaken here provided evidence suggesting that BMI has a broad causal impact on fecal microbiota variation. Effect estimates were derived from both observational linear modeling in cross-section and from a one-sample MR framework within 2,225 individuals from the Flemish Gut Flora Project (FGFP). Previous work has shown there to be very little evidence of population structure in FGFP [60]. Microbiota variation was modeled as five unique trait types: (1) abundance (AB), (2) presence absence (PA), (3) diversity (Div), (4) enterotype class (Entero) and (5) ratios (RA) of correlated abundance pairs, totaling 368 tested microbiota traits or MTs. Observational and MR effect estimates were strongly correlated with each other (Pearson’s r across all 368 traits = 0.69, p = 6.06×10^-54^; **Figure 5B**). All trait types exhibited both observational and causal associations with BMI. A total of 141 traits or 38% of those tested had an observational association with BMI (**Figure 2-4**), and a total of 41 MTs or 11% of those tested presented evidence to suggest that BMI causally influences their variation (**Table 2 and Figure 5-7**).

Previous work has provided evidence suggesting that the abundance of microbiota varies with BMI [86]. Of particular interest has been the relationship between phylum Firmicutes and phylum Bacteroidetes, the two most abundant phyla in gut microbiota (**Figure 4A**). It has been hypothesized that the abundance of Firmicutes would increase with increases in BMI and Bacteroidetes would decrease in a dependent fashion [38,39]. Similar to other work, we did not observe evidence that variation at either phylum associated with BMI individually (**Figure 4A**) or as a ratio [87]. However, at lower Firmicutes and Bacteroidetes taxonomic levels, we observed variation at numerous AB MTs to covary – in the form of RA MTs - within and between these phyla, such as with G_Bacteroides | G_Sporobacter or F_Lachnospiraceae | F_Porphyromonadaceae (**Figure 3 and 6**).

Association with cross-sectional increases in BMI were not consistent in direction (i.e., not exclusively positive or negative) within a phylum (**Figure 3 and 6**). We observed MTs, in both Firmicutes and Bacteroidetes phyla, to both increase and decrease in AB or PA with increases in BMI (**Figures 3 and 6**). This observation is particularly true for MTs in phylum Firmicutes, where MR estimates support observational estimates that yielded both positive and inverse associations (**Figure 7**). Specifically, G_Roseburia_0.988_AB of phylum Firmicutes had the strongest positive observational association among abundance traits (**Figure 2**) and its MR estimate was consistent with the observational data but did not meet our study’s multiple correction threshold to declare a causal relationship. While in contrast to observing little evidence for an association between BMI and phylum Firmicutes, this observation is, in general, consistent with previous work suggesting that increases in BMI increase the abundance in phylum Firmicutes [38,40,88,89]. However, in FGFP, phylum Firmicutes made up, on average, 60% (95% QI 26.8-82.4) of the fecal microbiota and *Roseburia* made up, on average, 14% (95% QI 0.4-42) of Firmicutes. Consequently, increases in *Roseburia* with increases in BMI would not appear to be enough, individually, to generate associations at the phylum level. This may be particularly true given that three other AB traits derived from three other microbiota taxa of phylum Firmicutes, including genera *Sporobacter*, have inverse associations with BMI (**Table 2**, **Figure 6**) and make up, on average, 6% (95% QI 0-26) of the read counts for Firmicutes, on average.

To the exclusion of unclassified MTs, eight genera-level MTs were associated with BMI in MR analyses (**Figure 7**) either individually (*Barnesiella*, *Sporobacter*) or as a product of their inclusion in a RA MT (*Alistipes*, *Bacteroides*, *Blautia*, *Clostridium IV*, *Roseburia* and *Oscillibacter*). These eight genera are intercorrelated with each other, with *Roseburia* and *Bacteroides* being the only pair exhibiting no correlation (**Supplementary Figure 9**). However, among the 28 RA MTs, that could be constructed among these eight MTs, only those that included *Barnesiella or Sporobacter* exhibited evidence of a causal effect of BMI (**Supplementary Figure 9**). Perhaps, this observation was a product of their individual effects. Resultantly, these two genera represent those with the best evidence of being influenced by BMI even in the context of a network of genera that covary with cross-sectional increases in BMI (**Supplementary Figure 9**).

These data are consistent with BMI (or the conditions that are measured in proxy by it) having an effect on microbial abundance variation within and across phyla in a correlated and inverse manner (i.e., as the abundance of certain taxa increases, abundance of other taxa decreases). An advantage to including MT ratios is that ratio traits are not influenced by inter-individual variation in microbial load [90,91]. As such, evidence here suggests that BMI is acting on each MT in concert, and that the covarying abundance of microbiota in RA MTs are not the product of the compositional nature of microbiota data. In addition, these data would suggest that associating higher level taxonomic units like phyla to phenotypes of interest, like BMI or irritable bowel syndrome, may not be instructive or even appropriate given the biological variability and complex interactions between microbiota and host within a phylum, or perhaps even at lower taxonomic levels, such as class.

The results presented here also suggest that sex-specific analyses can identify novel, sex-specific causal relationships. Notably, six MTs were identified to be causally influenced by BMI in females but not in males (**Supplementary Figure 7**). Most prominently, higher BMI appears to decrease the presence of genera *Aldercreuzia* (G_Aldercreuzia_0.66_PA), a taxon associated with phytoestrogen metabolism and equol production, which may have beneficial health effects (**Figure 6**) [92]. Additionally, two other MT that causally associated with BMI in females involved family Coriobacteriaceae (G_unclassified_F_Coriobacteriaceae_0.756_AB and F_Coriobacteriaceae | G_Ruminococcus2). Coriobacteriaceae belongs to the same bacterial class, Coriobacteriia, as *Aldercreuzia*, and is a family of bacteria known to include bacteria believed to also be phytoestrogen metabolizers [93]. Collectively, these findings highlight the potential utility of combining the general population and the sex-specific PGSs in sex-stratified MR analyses to identify causal relationships that may be specific to one sex.

This study had several limitations. It had a relatively small sample population of 2,225 middle aged individuals from the Flemish Gut Flora Project and was not followed up in an independent collection. The sample population is from the Flemish region of Belgium and individuals are of Northern European ancestry and, as such, inference made from results here may be limited to sample populations of similar ancestry and environments. We used rarefied 16S rRNA relative abundance data in our analyses and not absolute abundance. The outcome traits being studied are complex, with highly skewed distributions, necessitating the need for rank inverse normal transformation of the data prior to parametric analyses. This reduced the utility and meaning behind the effect estimates and complicates any translational science.

Interpretation of ratio traits are inherently complex, as they are derived from the ratio of two aforementioned complex and skewed distributions, which further complicates biological interpretation. Additionally, no non-linear analyses were performed, and if such relationships exist between BMI and microbiota trait variation then our linear modeling may fail to identify associations and/or effect estimates may be erroneous. This may be relevant as our sample population was relatively healthy, where just 13.4% of the sample had obesity and 44.5% had overweight or obesity at the time of sampling. Because we used linear models to estimate the effect of a rank-normalized standard deviation unit increase in BMI, rather than modeling BMI as a binary exposure (e.g., obese vs. non-obese, using a BMI ≥30 threshold), we may have been underpowered to detect associations that are non-linear or that emerge more abruptly at higher BMI levels. In such cases, binary traits could be more sensitive than continuous models, especially if no association exists below or above a certain threshold. Finally, the genetic instrument(s) used for BMI was highly polygenic and individual SNPs (or their LD neighbors) may influence BMI through diverse biological pathways, from behavior to energy storage. As a result, the MR effect estimates may represent some average effect across all such mechanisms that may be limiting causal inference.

In summary, the data presented here provide strong evidence that BMI has a causal impact on gut microbiome variation in humans. The mechanism by which BMI, itself a complex trait, impacts gut microbiome variation has yet to be identified but will certainly lie in altering host environment through numerous pathways including diet, physical activity, behavior, insulin sensitivity, and inflammation. Whilst it would be instructive to perform the reverse MR (i.e., using MTs as exposures), instrumenting the microbiome is presently not a viable option, as only one MT, namely *Bifidobacterium*, has been robustly associated with one genetic marker across multiple studies [60,62,64,66,94–99], namely the lactose tolerance allele at MCM6/LCT, and even its use requires careful considerations [60,61]. Future work may address this limitation as ever larger GWASs become available. At present, however, informative MR studies in this context are constrained by the lack of knowledge regarding how the few identified (but not replicated) genetic variants relate to other MTs, as well as the subsequent inability to apply appropriate sensitivity analyses (e.g., pleiotropy-robust methods) to test for violations of core MR assumptions. For example, the *ABO* locus is associated with certain MT(s) in several studies but is associated with different MTs in different populations [94,95] or none at all in others [100,101].

While it remains an open question if modulating the gut microbiome of individual humans can have a measurable influence on adiposity traits [102–104], or any health outcome, this work supports the inverse conclusion: changes in adiposity, here measured as cross-sectional increases in BMI, may well have a causal impact on gut microbiome variation. This impact spans multiple features including diversity, relative abundance, presence, enterotype assignment, and ratios of correlated taxa. These results suggest that observational studies linking microbiome features to adiposity should be reappraised in light of this likely direction of effect. Moreover, the specific agents driving apparent microbiome effects may not always be clear. Our findings suggest potential interactions between upstream factors captured by BMI, such as dietary behavior, and downstream modulators of microbiome variation, such as the dietary substrates available to gut microbes. Together, these insights highlight the need for integrative approaches that consider both host and microbial contributions to metabolic health.

## Supporting information

STROBE MR

## Data Availability

All data produced in the present study are available upon reasonable request to the the authors and the Flemish Gut Flora Project.

https://github.com/hughesevoanth/FGFP_BMI_MR

## Acknowledgements

We acknowledge the contribution of Flemish general practitioners and pharmacists to data and sample collection. Finally, we thank all FGFP volunteers for their participation in the project.

