## Supplementary Figures for "Estimating the causal effect of body mass index on gut microbiota variation"

### Manuscript

### Supplementary Figure 1: A flow diagram for defining microbial traits

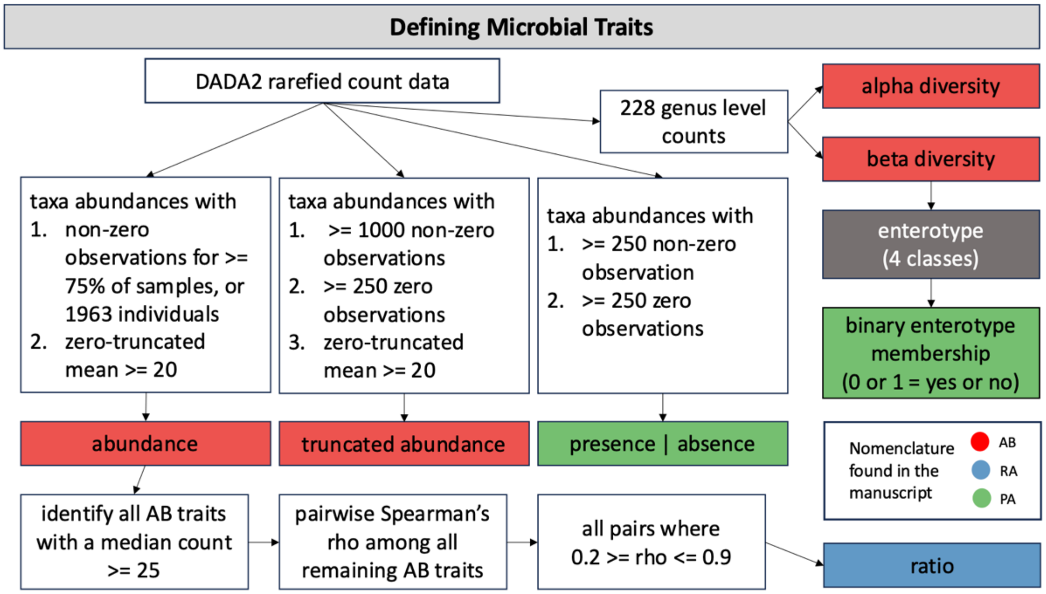


**Figure S1: A flow diagram for defining microbial traits:** The flow diagram provides information about how the microbial traits or MTs were derived. There are six specific types of traits. They are diversity, enterotype, abundance, (zero-)truncated abundance, ratios of abundances, and presence or absence traits. These represent two analytical categories, namely binary traits (presence|absence and the binary enterotypes (0|1 or yes|no a sample does or does not belong to an enterotype class)), and continuous traits (diversity, (zero-truncated) abundance, and ratios). Binary traits were modeled with logistic regression and all others were modeled assuming a normal distribution, after inverse rank normal transformation.

### Supplementary Figure 2: Correlation plot among all microbial traits

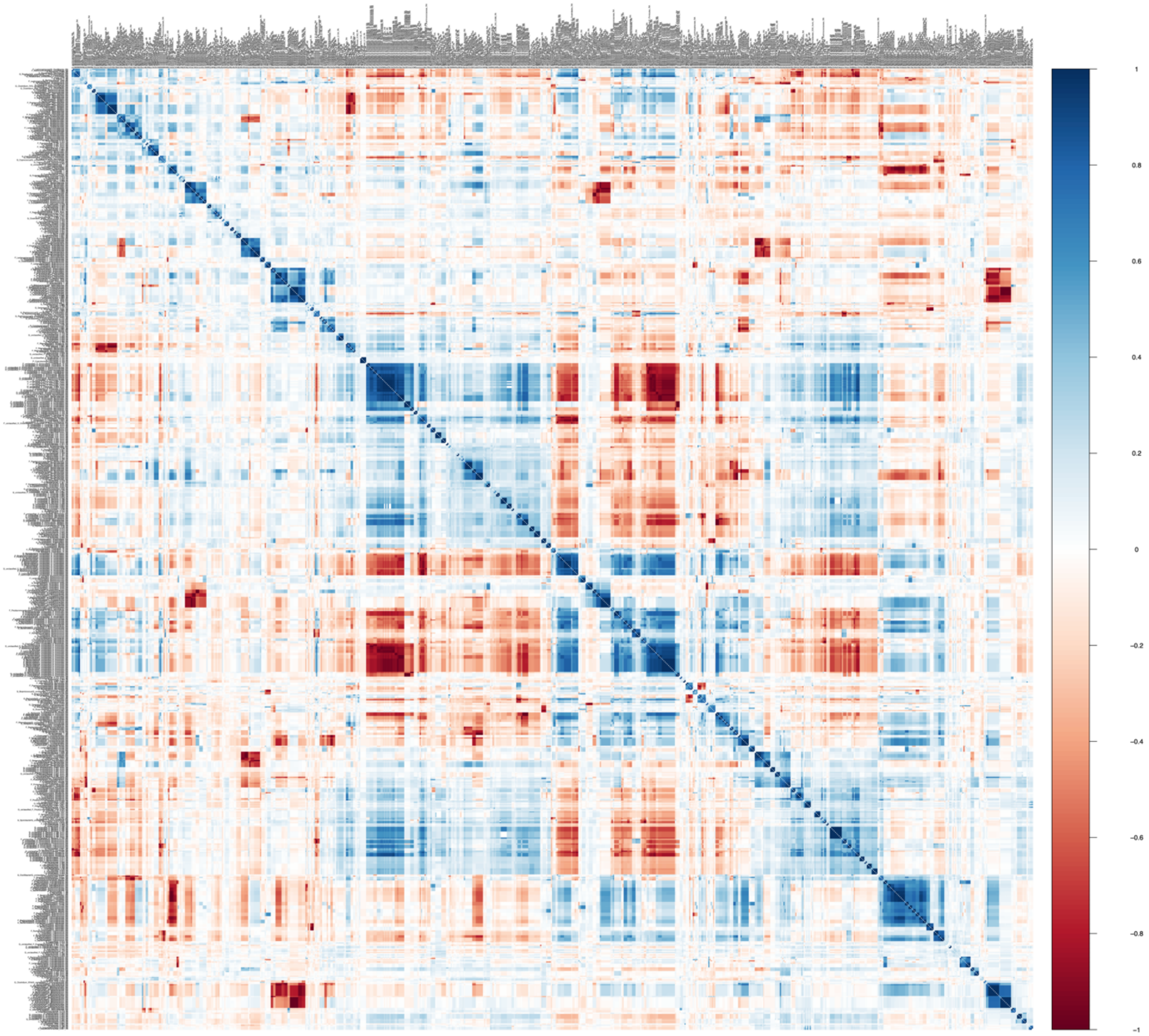


**Figure S2: Correlation plot among all identified microbial traits:** A correlation plot of Spearman's rho estimates from pairwise complete observations among abundance (AB), truncated abundance (trunc_AB), and present or absent (PA) traits identified in the FGFP DADA2 rarefied count data.

### Supplementary Figure 3: Correlation plot among non-redundant microbial traits

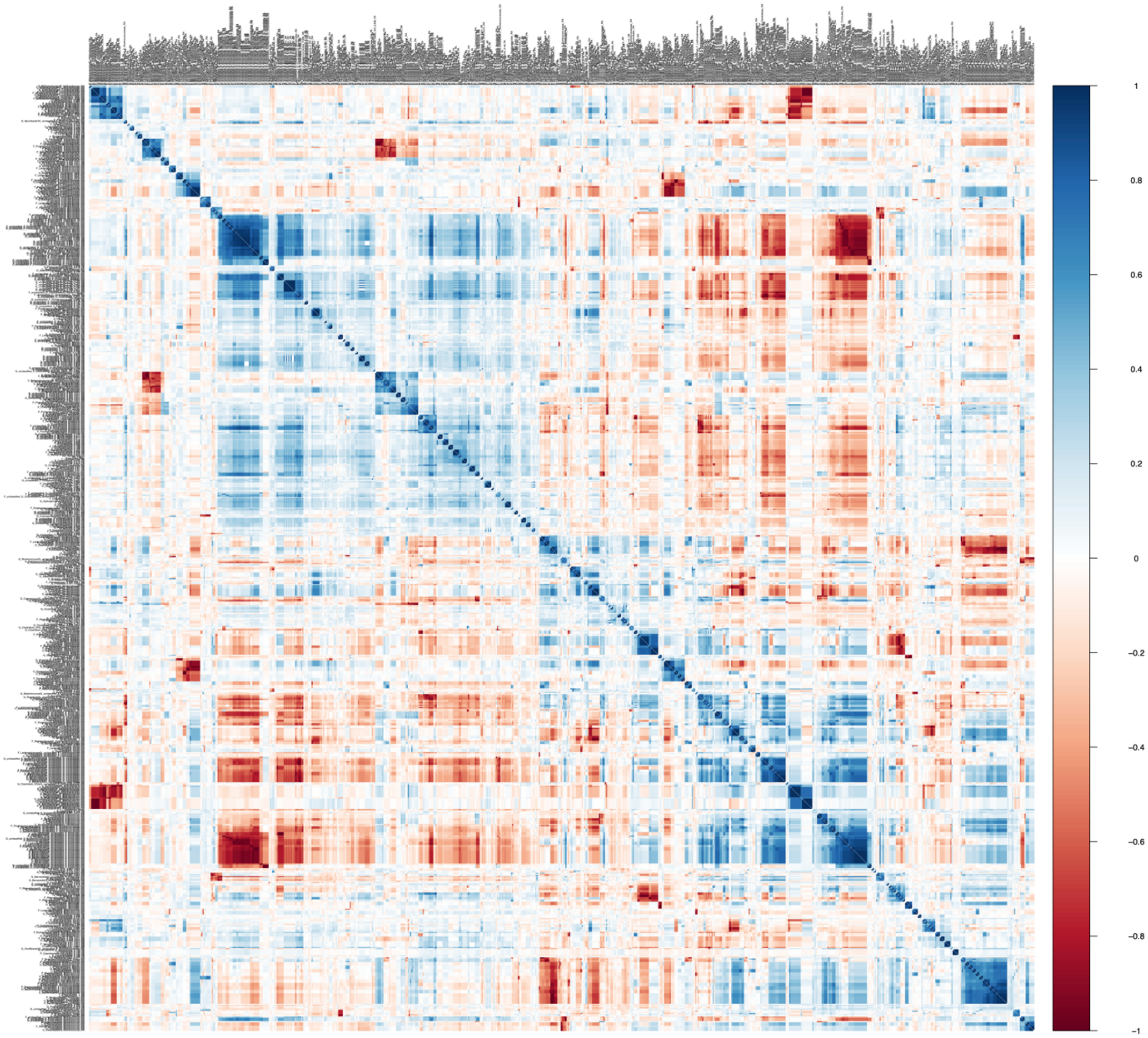

**Figure S3: Correlation plot among non-redundant microbial traits:** A correlation plot of Spearman's rho estimates from pairwise complete observations among abundance (AB), truncated abundance (trunc_AB), and present or absent (PA) traits identified in the FGFP DADA2 rarefied count data but filtered to exclude those that are fully redundant or had a Spearman's rho >= 0.99. Lower phylogentic MTs were retained while high level taxanomic units were removed. For example, P_Bacteroidetes, C_Bacteroidia, and O_Bacteroidales (Phylum, Class, Order) have the same count values (Figure S1) so P_Bacteroidetes, C_Bacteroidia were removed from further analysis while O_Bacteroidales was retained.

### Supplementary Figure 4: Principal Component Analysis Plot

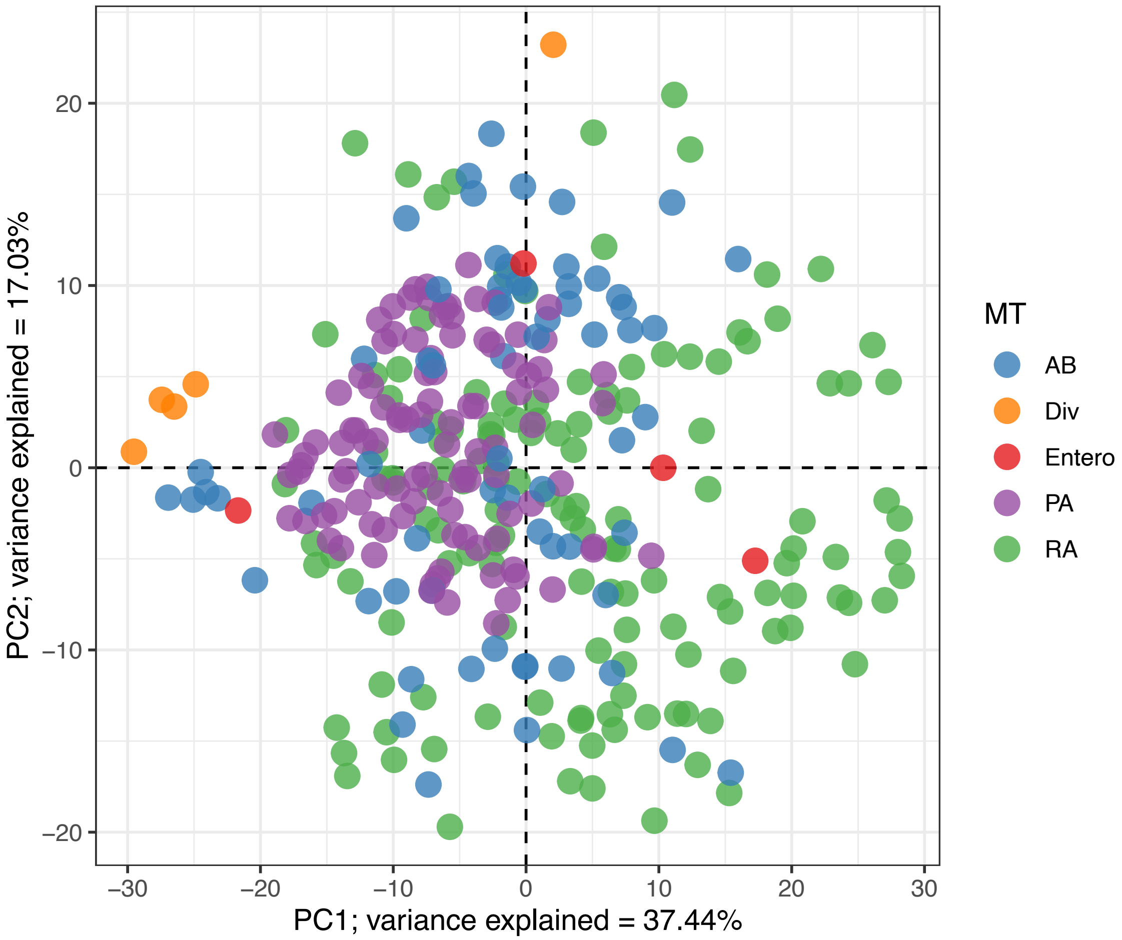


**Supplementary Figure 4: Principal component analysis plot.** A PCA plot of the 368 primary microbiota traits (MTs), derived from a Spearman's correlation matrix. Each dot represents a MT which are color coded by their trait type: blue for abundance traits (AB), orange for diversity traits (Div), red for enterotypes (Entero), purple for presence or absence traits (PA), and green for ratio traits (RA). The x- and y-axis denote the first and second principal component or eigenvector values for each MT, respectively. Also, presented on each axis is the proportion of total microbiota variance explained, or eigenvalues, by PC1 and PC2. We used the PCA eigenvalues to estimate the number of effective markers (Me) or microbial traits that were present in the data set. This was done by identifying the number of PCs need to sum up to at least 95% of the total variance. Here the estimate for Me was 33.

### Supplementary Figure 5: Analysis flow diagram

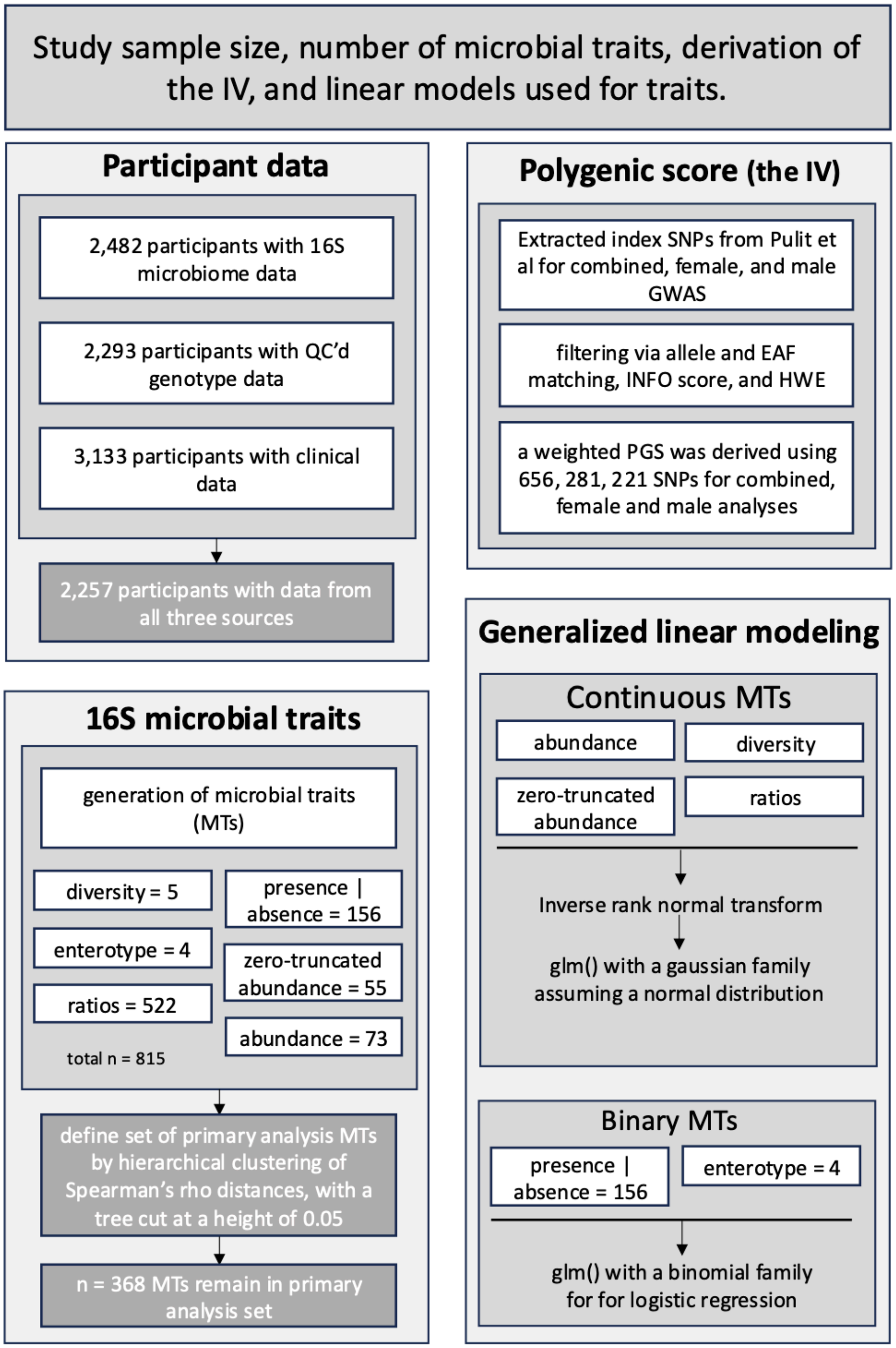


**Supplementary Figure 5: Analysis flow diagram.** The flow diagram provides information on the study data and analytical framework. The "Participant data" provides information on the sample size for each source of data and the overlap. The "16S microbial traits" defines the number of traits for each trait type in the study, as well as how the primary, non-redundant, study traits were identified. The "Polygenic Score" outlines the steps taken to generate the instrumental variable (IV). The "Generalized linear modeling" defines which traits were analyzed linear or logistic regression.

### Supplementary Figure 6: Scatterplot of general population and sex-specific MR estimates.

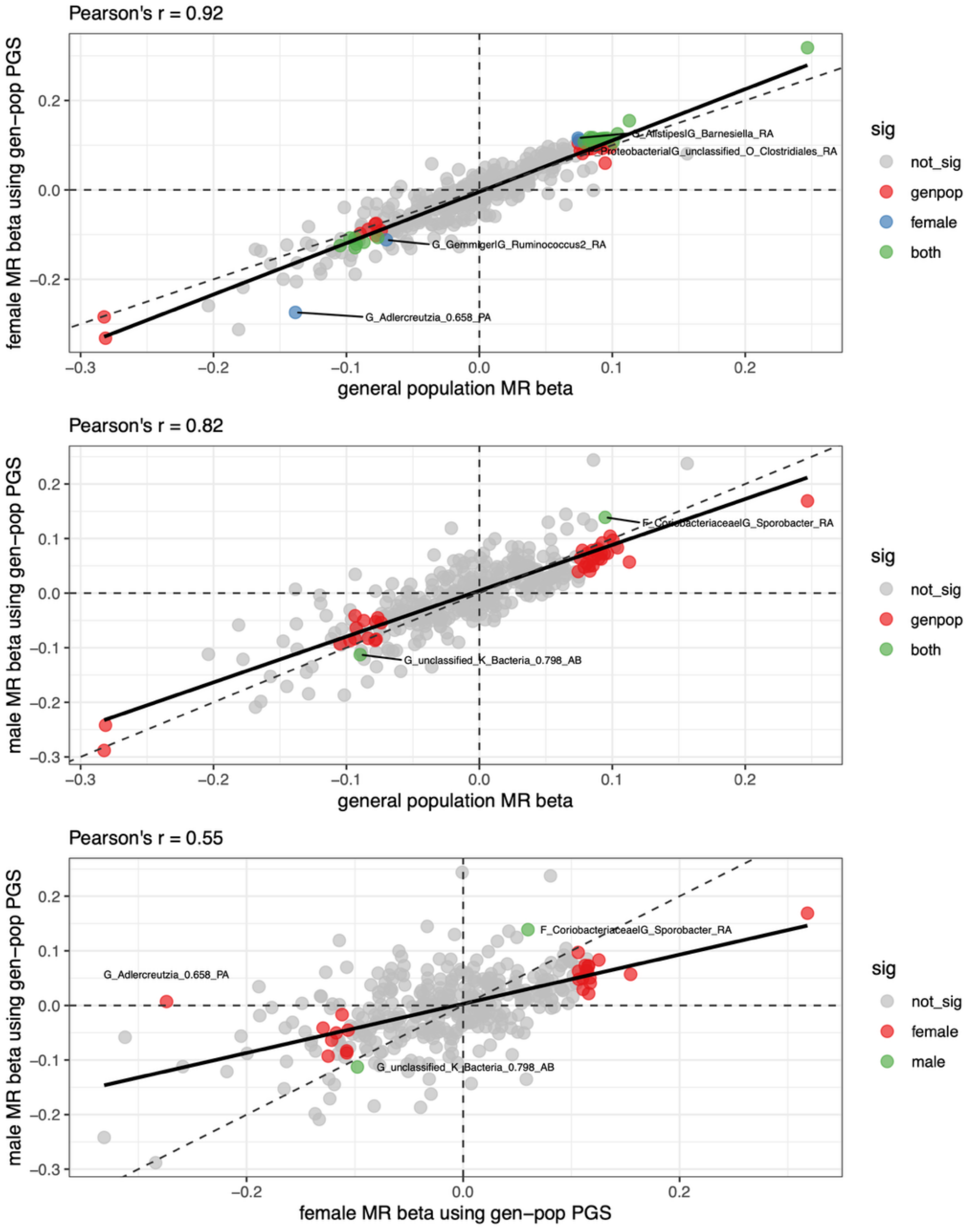


**Supplementary Figure 6: Scatterplot of general population and sex-specific MR estimates.** Scatterplot of MR effects. Each scatterplot presents a comparison of MR point estimates from two different MR models. In the title of each plot the estimated Pearson's correlation coefficient, r, is reported. Points in each figure are color coded to indicate their MR association signals. Points that are grey had no association in either analysis, while other colors define if there were associated in both – the x-axis and y-axis - analyses or in just one. The dashed horizontal line is the equivalency line, and the solid black line is the best fit line from a univariable linear regression of y on x. (TOP) Primary MR effect estimates from the sex-combined general population analysis is plotted on the x-axis, and female-specific MR estimates derived using the general population PGS as the instrument are plotted on the y-axis. (MIDDLE) Primary MR effect estimates from the sex-combined general population analysis is plotted on the x-axis, and male-specific MR estimates derived using the general population PGS as the instrument are plotted on the y-axis. (Bottom) Female-specific MR effect estimates when using the general population PGS as an instrument are plotted on the x-axis, and male-specific MR estimates derived when using the general population PGS as the instrument are plotted on the y-axis.

### Supplementary Figure 7: Forest plot of female specific MR association.

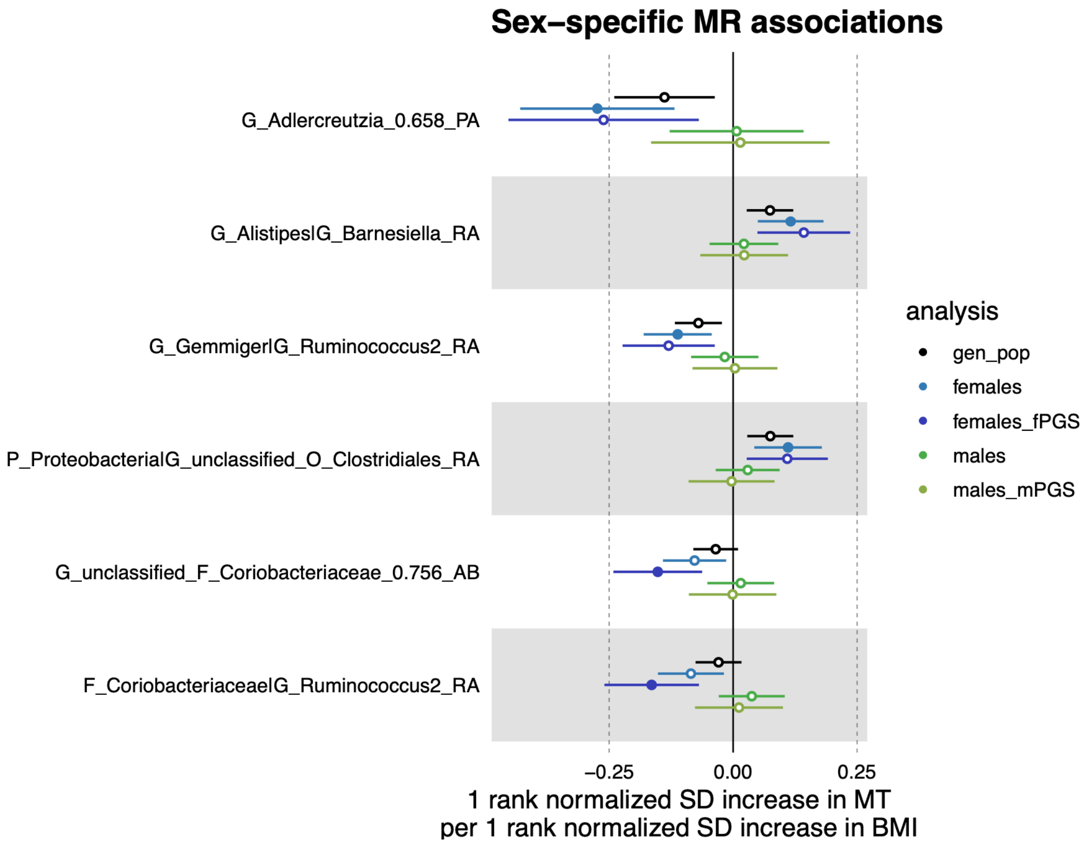


**Supplementary Figure 7: Forest plot of female specific MR associations.** MR effect estimates for BMI on MTs are illustrated as the dots along with their 95% confidence intervals. MR effect estimates for the general population (black), females using the general population PGS as an instrument (blue), females using the female-specific PGS as an instrument (dark blue), males using the general population PGS as an instrument (green), and males using the male-specific PGS as an instrument (dark green) are illustrated. Point estimates that are solid in color surpassed our multiple testing burden to declare an association (P>0.05/33), while those with a white dot did not. MTs are organized by their phylum or trait type.

### Supplementary Figure 8: Scatterplot of sex-specific MR estimates.

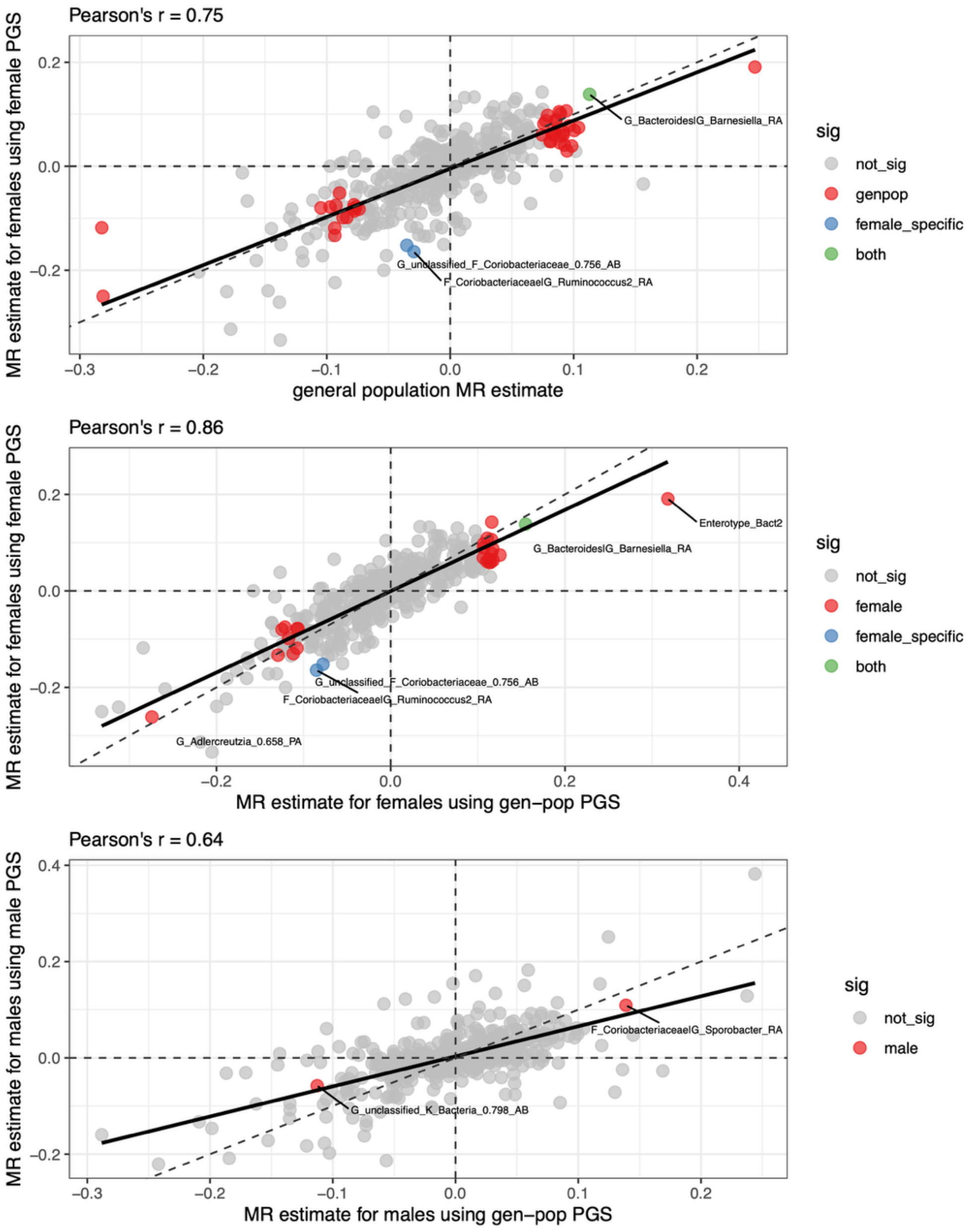


**Supplementary Figure 8: Scatterplot of sex-specific MR estimates.** Scatterplot of MR effects. Each scatterplot presents a comparison of MR point estimates from two different MR models. In the title of each plot the estimated Pearson's correlation coefficient, r, is reported. Points in each figure are color coded to indicate their MR association signals. Points that are grey had no association in either analysis, while other colors define if there were associated in both – the x-axis and y-axis - analyses or in just one. The dashed horizontal line is the equivalency line, and the solid black line is the best fit line from a univariable linear regression of y on x. (TOP) Primary MR effect estimates from the sex-combined general population analysis is plotted on the x-axis, and female-specific MR estimates derived using the female specific PGS as the instrument are plotted on the y-axis. (MIDDLE) Female-specific MR effect estimates when using the general population PGS as an instrument are plotted on the x-axis, and female-specific MR estimates derived when using the female specific PGS as the instrument are plotted on the y-axis. (Bottom) Male-specific MR effect estimates when using the general population PGS as an instrument are plotted on the x-axis, and male-specific MR estimates derived when using the female specific PGS as the instrument are plotted on the y-axis.

Supplementary Figure 9: Genera associated with BMI in MR analyses.

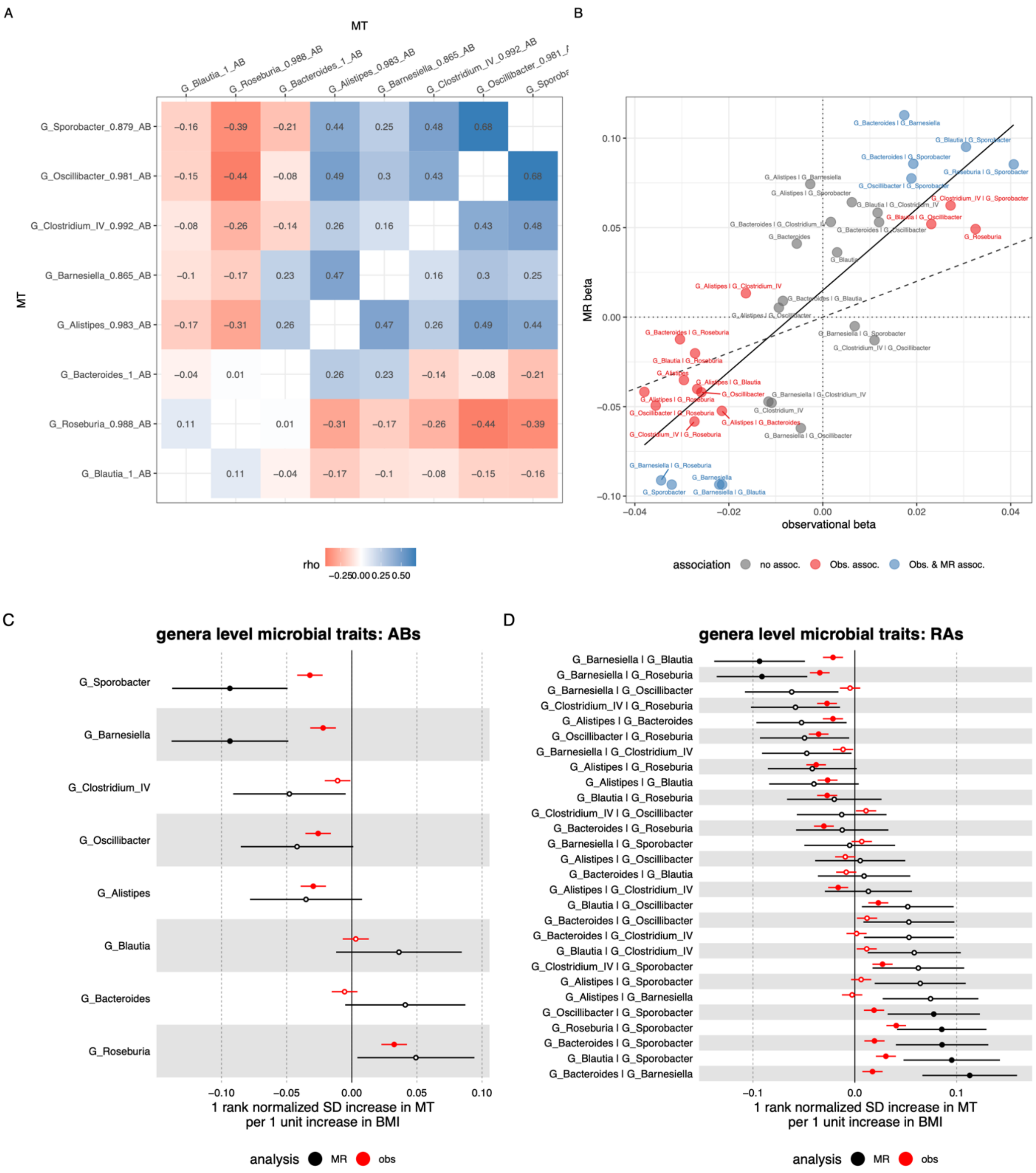


**Supplementary Figure 9: Genera associated with BMI in MR analyses.** (A) Spearman's rho among the eight genera associated with BMI ether individually or as observed in ratio traits, with Spearman's rho stated in each cell, blue cells indicating positive pairwise associations and red cells indicating negative correlations. The cells on the diagonal were removed as they are each self-correlations with a value of one. (B) The correlation structure among the observational and MR effect estimates, color coded to indicate those microbial traits with no association (grey), those associated with BMI in observational analyses (red), or those associated in both observation and MR analyses (blue). The solid line is a best fit line, while the dashed line is an equivalency (x=y) line. (C) A forest plot of observational (red) and MR (black) effect estimates and standard errors for the eight genera. (D) A forest plot of observational (red) and MR (black) effect estimates for each of the 28 ratio microbial traits one could make among the eight genera. Note that these ratio MTs may not be found in SupTables, as the ratio traits in this figure were made explicitly for this figure - for the purposes of exploration and completeness.
