## Supplementary material for "Estimating the causal effect of body mass index on gut microbiota variation": STROBE MR

**STROBE-MR checklist of recommended items to address in reports of Mendelian randomization studies**[1 2]

| Item No. | Section | Checklist item | Page No. | Relevant text from manuscript |
|---|---|---|---|---|
| 1 | **TITLE and ABSTRACT** | Indicate Mendelian randomization (MR) as the study's design in the title and/or the abstract if that is a main purpose of the study | 1 | We used fecal 16S rRNA data from 2,225 individuals from the Flemish Gut Flora project to derive 368 microbiota traits (MTs) including diversity, abundance, presence, or absence, enterotype class and abundance ratio. We assessed the relationship between BMI and those traits in observational and Mendelian randomization (MR) frameworks, the latter being used to estimate a causal effect of BMI on MT variation. |
| | **INTRODUCTION** | | | |
| 2 | **Background** | Explain the scientific background and rationale for the reported study. What is the exposure? Is a potential causal relationship between exposure and outcome plausible? Justify why MR is a helpful method to address the study question | 2-3 | "Cross-sectional increases in BMI have been associated with a variety of diseases including cardiometabolic disease, cardiovascular disease and type 2 diabetes, as well as with some cancers and decreased life expectancy."<br>and<br>"Numerous studies have illustrated that gut microbiota are associated with environmental and host physiology variables. Notably, these associations extend to variation in BMI and a wide range of diseases, including metabolic disorders such as type 2 diabetes, inflammatory bowel syndrome, Crohn's disease, neurological traits such as Parkinson's disease and stress, as well as cancers and other complex traits."<br>and<br>"Common difficulties with observational analysis of variation in gut microbiota and disease traits are confounding and reverse causality. Specifically, distinguishing whether variations in gut microbiota precede and influence disease risk, are caused by the disease itself, or simply coincide with disease |

| | | | | |
|---|---|---|---|---|
| | | | | due to shared confounders (such as diet, lifestyle, or genotype) is difficult to ascertain without well-executed prospective studies and randomized controlled trials. In lieu of these studies, Mendelian randomization (MR) may aid assertions of causality in studies of microbiome and BMI."<br>and<br>"In this context, rather than considering the gut microbiota as an exposure, it is, here, modelled as a response variable, specifically within a MR framework (**Figure 1**)." |
| 3 | **Objectives** | State specific objectives clearly, including pre-specified causal hypotheses (if any). State that MR is a method that, under specific assumptions, intends to estimate causal effects | 3 | "We aimed to provide two estimates of the association between BMI and gut microbiota variation. First, human genetic and 16S rRNA gut microbiota data from the Flemish Gut Flora Project (FGFP) were used to estimate a conventional cross-sectional estimate of the effect of BMI on variation in fecal microbiota. Second, a within-sample or one-sample MR framework was used to obtain a causal effect estimate of BMI on variation in gut microbiota. Analyses were conducted in a sex-combined and sex-stratified analyses – motivated by the dimorphism in adiposity distributions among the sexes, the large associated contribution of sex to variation in the gut microbiota, and variation in disease risk among the sexes." |
| | **METHODS** | | | |
| 4 | **Study design and data sources** | Present key elements of the study design early in the article. Consider including a table listing sources of data for all phases of the study. For each data source contributing to the analysis, describe the following: | | |
| | a) | Setting: Describe the study design and the underlying population, if possible. Describe the setting, locations, and relevant dates, including periods of recruitment, exposure, follow-up, and data collection, when available. | 4 | "The study sample population was from the Flemish Gut Flora Project (FGFP), a population-based study in the Flanders region of Belgium focused on gut flora and health[1]. Sampling procedures are described in full elsewhere. Volunteers were invited to participate through local media and provided |

| | | | |
|---|---|---|---|
| | | | informed consent by mail. … Between June 2013 and April 2016 medical questionnaires were completed by each volunteer's general practitioner who also recorded anthropomorphic and basic clinical measurements. Stool samples were collected remotely with sampling kits sent to volunteer homes. Samples were collected and stored by volunteers at -18°C. Upon arrival at the research facility, they were stored again at -18°C until collection and long-term storage at -80°C." |
| b) | Participants: Give the eligibility criteria, and the sources and methods of selection of participants. Report the sample size, and whether any power or sample size calculations were carried out prior to the main analysis | 9 | "After combining genotype data (PGS), microbiome traits, anthropomorphic and clinical variables the study data set consisted of 2,257 individuals (**Supplementary Figure 5**). However, after accounting for missingness at covariables used in association analyses the maximum sample size in any single linear model was 2,225." |
| c) | Describe measurement, quality control and selection of genetic variants | 7-8 | "Genotype data from a total of 2,293 FGFP volunteers were included in the study and a complete description of genotyping and quality control is described in detail in Hughes et al. In brief, 2,646 individuals were genotyped on Illumina Human Core Exome array (v1.0 and v1.1), processed on GenomeStudio v.2.0.4 and quality controlled to remove unmapped variants, duplicated sites, those with missingness greater than 5%, and Hardy–Weinberg equilibrium deviation $P<1\times10^{-5}$. Quality control was then performed on samples to remove individuals with array failures, variant missingness greater than 5%, excess heterozygosity and cryptic relatedness. Genotype data was then merged with all Phase 3 data of the 1000 Genomes data set to identify individuals of European genetic ancestry using principal component analysis. Following these quality control steps 2,293 individuals and 509,886 variants remained. Individuals were then imputed to a version of the Haplotype Reference Consortium |

| | | | | | |
|---|---|---|---|---|---|
| | | | | | that included the UK10K and all individuals from Phase 3 of the 1000 Genomes Project."<br>and<br>"The instrumental variable for BMI used in this study is a polygenic score (PGS). We are explicitly not referring to the PGS as a risk score as BMI itself is not a disease. The PGS was derived from a meta-analysis of genome-wide association studies (GWASs) that was conducted in a sex-combined and (self-reported) sex-specific manner [68]. In total, three PGS instrumental variables were constructed: one for a general European population, one for European females and one for European males." |
| | | d) | For each exposure, outcome, and other relevant variables, describe methods of assessment and diagnostic criteria for diseases | 4-7 | Section "Defining Microbial Traits" and "Construction of the polygenic score" |
| | | e) | Provide details of ethics committee approval and participant informed consent, if relevant | 4 | "FGFP procedures were approved by the Medical Ethics Committee of the University of Brussels/Brussels University Hospital (approval no. 143201215505, 5 December 2012) and a declaration concerning the FGFP privacy policy was submitted to the Belgian Commission for the Protection of Privacy." |
| 5 | **Assumptions** | | Explicitly state the three core IV assumptions for the main analysis (relevance, independence and exclusion restriction) as well assumptions for any additional or sensitivity analysis | 7 | Section "MR assumptions"<br>"With all MR analyses there are three core assumptions (**Figure 1**). First, the relevance assumption dictates that the instrument(s) must be robustly associated with the exposure. Second, the independence assumption states that the instrument(s) must not associated with confounders associated to the outcome. Third, the exclusion restriction assumption states that the instrument is only related to the outcome via the exposure, that is, there is no horizontal pleiotropy or path from the instrument to the outcome except via the exposure." |

| | | | | |
|---|---|---|---|---|
| 6 | **Statistical methods: main analysis** | Describe statistical methods and statistics used | | |
| | a) | Describe how quantitative variables were handled in the analyses (i.e., scale, units, model) | 10 | "Given that the exposure trait – in its published GWAS - and all outcome abundance traits were rank-based inverse normal transformed prior to running linear models all effect estimates for AB, RA and diversity traits represent a rank normal standard deviation unit of change for each rank normal standard deviation unit increase in BMI. Effect estimates for binary PA traits are presented in log-odds, such that they represent the log-odds of presence (or being assigned to Enterotype class 'X'; coded as 1=Enterotype 'X', 0=other) for each rank normal standard deviation unit increase in BMI." |
| | b) | Describe how genetic variants were handled in the analyses and, if applicable, how their weights were selected | 8 | Section "Construction of a BMI polygenic score"<br>"The combined, female, and male PGS were then constructed by weighting each effect allele by the (respective combined, female, and male) effect estimates provided by Pulit *et al.* and summing across all weighted effect alleles carried by an individual." |
| | c) | Describe the MR estimator (e.g. two-stage least squares, Wald ratio) and related statistics. Detail the included covariates and, in case of two-sample MR, whether the same covariate set was used for adjustment in the two samples | 10 | "One-sample MR analyses were then performed to estimate the causal effect of BMI on each MT (**Figure 1**). To do so, stage one of the two-stage least squares (MR) model was run by defining BMI as the outcome, the PGS as the exposure and including sex and age as covariates – again using the glm() function. Stage two of the two-stage least squares (MR) model was then estimated using the ivglm() function of the 'ivtools' R package using the estimation method "ts" or two-stage which derives the causal effect estimates and standard errors. The ivglm() function takes as input the stage one model and the observational model described above. A Breusch-Pagan test, Wald Test, Wu-Hausman endogeneity test, Type II ANOVA and $\eta^2$ statistics were then run and derived on each MR model. All |

| | | | | |
|---|---|---|---|---|
| | | | | observational and MR analyses described in this section were run with the bespoke wrapper function ivtoolsfit() that can be found in the study's GitHub repository." |
| | | d) Explain how missing data were addressed | 9 | "After combining genotype data (PGS), microbiome traits, anthropomorphic and clinical variables the study data set consisted of 2,257 individuals (**Supplementary Figure 5**). However, after accounting for missingness at covariables used in association analyses the maximum sample size in any single linear model was 2,225." |
| | | e) If applicable, indicate how multiple testing was addressed | 6-7 | Section "Effective number of microbial traits"<br>"To determine the effective or independent number of MTs that remain in this inter-correlated data set (n = 368, **Supplementary Figure 3**), we followed the procedure of Gao *et al*. [79]. First a Spearman's correlation matrix was generated among all MTs in the primary data set. Second, a principal component analysis was performed using this correlation matrix, which allows for data missingness and has the advantage of not requiring a data transformation given the use of a ranked correlation (**Supplementary Table 2, Supplementary Figure 4**). Third, we identified the number of components needed to explain at least 95% of the total variance in the data set. From this, we estimated that there are 33 effectively independent MTs in the data set. This estimate of the effective number of traits was then used to derive a study-wide data-reduced Bonferroni p-value threshold of 0.05/33 or $1.515 \times 10^{-3}$." |
| 7 | **Assessment of assumptions** | Describe any methods or prior knowledge used to assess the assumptions or justify their validity | 8 | MR Assumption 1: "In total, three PGS instrumental variables were constructed, one for a general European population, one for European females and one for European males. We used index variants from Pulit et al (taken from Pulit *et al.* **Supplementary Table 1**), which were those defined as being associated with BMI via |

| | | | | |
|---|---|---|---|---|
| | | | | conditional and joint association analysis at $P<5\times10^{-9}$ by the authors."<br>and<br>Assumption that BMI influences outcome: Introduction paragraph two. |
| 8 | **Sensitivity analyses and additional analyses** | Describe any sensitivity analyses or additional analyses performed (e.g. comparison of effect estimates from different approaches, independent replication, bias analytic techniques, validation of instruments, simulations) | 10 | "For sensitivity analyses, we repeated the observational and MR analyses as described above but include 16S total read count (the number of sequencing reads prior to being rarefied), Bristol stool score, J01 antibiotic use (yes = 1, no = 0), smoking status (never, ever, current), and household monthly income as additional model covariates (**Table 1**). Total read counts, Bristol stool score, and antibiotic use have known large effects on our outcome and are being included as precision variables to reduce technical or measurement error in both observational and MR analyses and to theoretically assess possible confounding in the observational analyses. Smoking status and household income, in contrast, are known to associate with our exposure and may be possible confounders of our exposure-outcome relationship in observational analyses." |
| 9 | **Software and pre-registration** | | | |
| | a) | Name statistical software and package(s), including version and settings used | 11 | "All statistical analysis were conducted in the R language (v 4.0.2, Taking Off Again) and all bespoke functions and analytical code for the study can be found in the GitHub repository https://github.com/hughesevoanth/FGFP_BMI_MR. " |
| | b) | State whether the study protocol and details were pre-registered (as well as when and where) | | NA |
| | **RESULTS** | | | |
| 10 | **Descriptive data** | | | |

| | | | | |
|---|---|---|---|---|
| | a) | Report the numbers of individuals at each stage of included studies and reasons for exclusion. Consider use of a flow diagram | 9 | "After combining genotype data (PGS), microbiome traits, anthropomorphic and clinical variables the study data set consisted of 2,257 individuals (**Supplementary Figure 5**). However, after accounting for missingness at covariables used in association analyses the maximum sample size in any single linear model was 2,225."<br>and<br>Flow diagram in Sup Figure 5<br>And<br>The 'n' for each individual regression is report in the Supplementary Tables 3, 4, and 5 |
| | b) | Report summary statistics for phenotypic exposure(s), outcome(s), and other relevant variables (e.g. means, SDs, proportions) | | Table 1 reports BMI<br>Supplementary Table 1 reports summary statistics on all outcome traits. |
| | c) | If the data sources include meta-analyses of previous studies, provide the assessments of heterogeneity across these studies | | NA |
| | d) | For two-sample MR:<br>i. Provide justification of the similarity of the genetic variant-exposure associations between the exposure and outcome samples<br>ii. Provide information on the number of individuals who overlap between the exposure and outcome studies | | NA |
| 11 | **Main results** | | | |
| | a) | Report the associations between genetic variant and exposure, and between genetic variant and outcome, preferably on an interpretable scale | | Supplementary Table 3, 4, and 5 |
| | b) | Report MR estimates of the relationship between exposure and outcome, and the measures of uncertainty from the MR analysis, on an interpretable scale, such as odds ratio or relative risk per SD difference | | Supplementary Table 3, 4, and 5 |
| | c) | If relevant, consider translating estimates of relative risk into absolute risk for a meaningful time period | | NA |
| | d) | Consider plots to visualize results (e.g. forest plot, scatterplot of associations between genetic variants and outcome versus between genetic variants and exposure) | | Figure 2,3, 5, and 6 |

| | | | | | |
|---|---|---|---|---|---|
| 12 | **Assessment of assumptions** | | | | |
| | | a) | Report the assessment of the validity of the assumptions | | NA |
| | | b) | Report any additional statistics (e.g., assessments of heterogeneity across genetic variants, such as $I^2$, Q statistic or E-value) | | NA |
| 13 | **Sensitivity analyses and additional analyses** | | | | |
| | | a) | Report any sensitivity analyses to assess the robustness of the main results to violations of the assumptions | 17 | Section "Sensitivity Analyses" and Supplementary Table 4 |
| | | b) | Report results from other sensitivity analyses or additional analyses | 18 | Section: "Untransformed analyses" and Supplementary Table 5 |
| | | c) | Report any assessment of direction of causal relationship (e.g., bidirectional MR) | | NA |
| | | d) | When relevant, report and compare with estimates from non-MR analyses | | Figure 6B illustrates relationship between all observational and MR estimates |
| | | e) | Consider additional plots to visualize results (e.g., leave-one-out analyses) | | NA |
| | **DISCUSSION** | | | | |
| 14 | **Key results** | | Summarize key results with reference to study objectives | 18 | "Analyses undertaken here provide evidence suggesting that BMI has a broad causal impact on fecal microbiota variation" |
| 15 | **Limitations** | | Discuss limitations of the study, taking into account the validity of the IV assumptions, other sources of potential bias, and imprecision. Discuss both direction and magnitude of any potential bias and any efforts to address them | 20 | "This study had several limitations. It had a relatively small sample population of 2,225 middle aged individuals from the Flemish Gut Flora Project and was not followed up in an independent collection. The sample population is from the Flemish region of Belgium and individuals are of Northern European ancestry and, as such, inference made from results here may be limited to sample populations of similar ancestry and environments. We used rarefied 16S rRNA relative abundance data in our analyses and not absolute abundance. The outcome traits being studied are complex, with |

| | | | | |
|---|---|---|---|---|
| | | | | highly skewed distributions, necessitating the need for rank inverse normal transformation of the data prior to parametric analyses. This reduced the utility and meaning behind the effect estimates and complicates any translational science.” |
| 16 | **Interpretation** | | | |
| | | a) Meaning: Give a cautious overall interpretation of results in the context of their limitations and in comparison with other studies | 21 | “In summary, the data presented here provide strong evidence that BMI has a causal impact on gut microbiome variation in humans. The mechanism by which BMI, itself a complex trait, impacts gut microbiome variation has yet to be identified but will certainly lie in altering host environment through numerous pathways including diet, physical activity, behavior, insulin sensitivity, and inflammation. Whilst it would be instructive to perform the reverse MR (i.e., using MTs as exposures), instrumenting the microbiome is presently not a viable option, as only one MT, namely *Bifidobacterium*, has been robustly associated with one genetic marker across multiple studies [60,62,64,66,94–99], namely the lactose tolerance allele at MCM6/LCT, and even its use requires careful considerations. Future work may address this limitation as ever larger GWASs become available. At present, however, informative MR studies in this context are constrained by the lack of knowledge regarding how the few identified (but not replicated) genetic variants relate to other MTs, as well as the subsequent inability to apply appropriate sensitivity analyses (e.g., pleiotropy-robust methods) to test for violations of core MR assumptions. For example, the *ABO* locus is associated with certain MT(s) in several studies but is associated with different MTs in different populations or none at all in others.” |
| | | b) Mechanism: Discuss underlying biological mechanisms that could drive a potential causal relationship between the investigated exposure and the outcome, and whether the gene-environment equivalence assumption is | 21 | The mechanism by which BMI, itself a complex trait, impacts gut microbiome variation has yet to be identified but will certainly lie in altering host |

| | | | | |
|---|---|---|---|---|
| | | reasonable. Use causal language carefully, clarifying that IV estimates may provide causal effects only under certain assumptions | | environment through numerous pathways including diet, physical activity, behavior, insulin sensitivity, and inflammation. |
| | | c) Clinical relevance: Discuss whether the results have clinical or public policy relevance, and to what extent they inform effect sizes of possible interventions | 21 | Same as statement below: |
| 17 | **Generalizability** | Discuss the generalizability of the study results (a) to other populations, (b) across other exposure periods/timings, and (c) across other levels of exposure | 21 | Last paragraph:<br>"While it remains an open question if modulating the gut microbiome of individual humans can have a measurable influence on adiposity traits [102–104], or any health outcome, this work supports the inverse conclusion: changes in adiposity, here measured as cross-sectional increases in BMI, may well have a causal impact on gut microbiome variation. This impact spans multiple features including diversity, relative abundance, presence, enterotype assignment, and ratios of correlated taxa. These results suggest that observational studies linking microbiome features to adiposity should be reappraised in light of this likely direction of effect. Moreover, the specific agents driving apparent microbiome effects may not always be clear. Our findings suggest potential interactions between upstream factors captured by BMI, such as dietary behavior, and downstream modulators of microbiome variation, such as the dietary substrates available to gut microbes. Together, these insights highlight the need for integrative approaches that consider both host and microbial contributions to metabolic health. " |
| | **OTHER INFORMATION** | | | |
| 18 | **Funding** | Describe sources of funding and the role of funders in the present study and, if applicable, sources of funding for the databases and original study or studies on which the present study is based | 22 | Section called "Funding" |
| 19 | **Data and data sharing** | Provide the data used to perform all analyses or report where and how the data can be accessed, and reference these sources in the article. Provide the statistical code needed to reproduce the results in the article, or report whether the code is publicly accessible and if so, where | 10 | Section "Code and data availability" |

| | | |
|---|---|---|
| 20 | **Conflicts of Interest** | All authors should declare all potential conflicts of interest |

This checklist is copyrighted by the Equator Network under the Creative Commons Attribution 3.0 Unported (CC BY 3.0) license.

1. Skrivankova VW, Richmond RC, Woolf BAR, Yarmolinsky J, Davies NM, Swanson SA, et al. Strengthening the Reporting of Observational Studies in Epidemiology using Mendelian Randomization (STROBE-MR) Statement. JAMA. 2021;under review.

2. Skrivankova VW, Richmond RC, Woolf BAR, Davies NM, Swanson SA, VanderWeele TJ, et al. Strengthening the Reporting of Observational Studies in Epidemiology using Mendelian Randomisation (STROBE-MR): Explanation and Elaboration. BMJ. 2021;375:n2233.
